# Early DNA methylation at the NGFI-A binding site of the *NR3C1* 1F promoter predicts cognitive functions at age five: evidence from the Parents as Teachers intervention in the ZEPPELIN study

**DOI:** 10.64898/2026.02.22.26346845

**Authors:** Elena Silvia Gardini, Alex Neuhauser, Simone Schaub, Isabelle Kalkusch, Patsawee Rodcharoen, Ulrike Ehlert, Andrea Lanfranchi, Gustavo Turecki, Peter Klaver

**Author notes:** Corresponding author (EG).

## Abstract

**Background:** The present study examines the link between DNA methylation at the nerve growth factor-induced protein A (NGFI-A) binding domain of the *NR3C1* 1F promoter and later cognitive functions in children from families living in disadvantaged psychosocial conditions.

**Methods:** Participants were 132 children who took part in a Swiss Parents as Teachers (PAT) randomized controlled trial (72 in the intervention group, 60 in the control group). DNA methylation was quantified from saliva samples collected at age three using sodium bisulfite next-generation sequencing (NGS). Cognitive functions were assessed at age five using the SON-R 2.5–7 Intelligence Test.

**Results:** (a) DNA methylation at age three was associated with lower IQ at age five through increased concentration problems (indirect effect = −0.083, p = .012); (b) parental disagreement was associated with concentration difficulties through DNA methylation (indirect effect = 0.064, p = .023); and (c) participation in the three-year PAT program was associated with lower methylation levels (β = −0.375, p = .041). The serial pathway from parental disagreement to IQ via methylation and concentration was not significant.

**Conclusions:** These findings provide preliminary evidence for an association between early DNA methylation at the NGFI-A binding site of the NR3C1 1F promoter and later cognitive functions in children and highlight the potential role of early life stressors and the PAT intervention in shaping these associations. However, given the exploratory nature of this study, replication in larger independent samples is warranted.

**Data Availability Statement:** All data supporting the findings of this study are publicly available on the SWISSUbase repository at https://doi.org/10.48573/h189-pd12 (DOI: 10.48573/h189-pd12). The specific data file used is named ‘NGFI-A binding site-m and cognition’.

## Introduction

Children growing up in families affected by psychosocial risks such as social isolation, poverty, or parental psychological distress are more likely to follow less favorable developmental and life-course trajectories, including lower academic achievement and socioeconomic status (SES) in adulthood (1,2).

Stress experienced by children living in psychosocially disadvantaged families is a key factor explaining these disparities (1,3). Psychosocial interventions aiming to strengthen parenting skills and improve the quality of the home learning environment are thought to reduce excessive stress experienced by the parents and children. Moreover, such interventions have been shown to confer substantial benefits by improving children’s cognitive development, school readiness, and the regulation of stress-related hormonal patterns (4–6).

At the biological level, psychosocial stressors stimulate the hypothalamic-pituitary-adrenal (HPA) axis, leading to the secretion of cortisol, which orchestrates physiological, cognitive, and behavioral changes that enable the organism to cope with challenging conditions (7). However, exposure to prolonged psychosocial stressors causes the HPA axis to adapt, leading to alterations in the stress response (7). Such alterations have been described in children exposed to disadvantaged psychosocial conditions and associated with impairments in attention regulation, cognitive functioning, and mental health (8–14). For example, based on findings that children with history of physical abuse showed lower regulatory responses to anger employment (15), Blair and Raver (8) suggested that the effect of child adversity on self-regulation development is mediated by stress hormones and neural connectivity. Attentional self-regulation in turn is a strong predictor of cognitive ability and school adjustment (see, for example (16,17)).

The molecular mechanism underlying the adaptation of the HPA axis in response to psychosocial stress involves DNA methylation (18). DNA methylation is an epigenetic mechanism that regulates gene expression through the binding of methyl groups at cytosines in cytosine–guanine dinucleotides (CpGs) (19). The termination of the HPA axis stress response is mediated by the binding of cortisol to the glucocorticoid receptor (GR) in the hypothalamus (20). Specifically, after binding to cortisol, the GR migrates in the cell nucleus, where it inhibits the expression of the corticotrophin releasing hormone (CRH) and consequently the releasing of cortisol from the adrenal cortex (20). Increased methylation of the GR gene (*NR3C1*) typically leads to decreased GR gene expression and therefore to an impaired termination of the stress response (21,22). In addition, lower levels of GR compromise the regulation of other various genes in the brain (23). Animal and human studies have shown that these altered physiological conditions negatively impact neurogenesis, synaptic plasticity and transmission in various brain regions such as the hippocampus, prefrontal cortex, and amygdala, leading to cognitive deficits, memory and learning dysfunctions, as well as vulnerability to psychiatric disorders (24–27).

A specific region in the exon 1F of the *NR3C1*, the nerve growth factor-induced protein A (NGFI-A) binding domain, has been shown to be particularly sensitive to increased methylation following early exposure to psychosocial stress (28–31). Reduced GR expression and altered HPA axis consequent to increased methylation in this DNA region have also been extensively reported (28–34). The mechanism underlying the association between psychosocial stress and methylation at the NGFI-A binding domain has been first described in 2004 in animal models (32). In this study, early stress in the form of low maternal care, such as low licking and grooming, decreased serotonin signaling in the hippocampus, leading to reduced expression of the immediate early gene NGFI-A (32). As described by Weaver et al. (35), reduced NGFI-A in early life failed to protect the *NR3C1* NGFI-A binding domain from methyl transferase (DNMT) activity, leading to increased methylation and other epigenetic modifications interfering with the GR gene transcription.

Because epigenetic patterns shaped during early life tend to persist, several authors have emphasized the importance of intervening during the early stages of development to counteract the potential long-term effects of early stress (1). Building on this concept, Gardini et al. (36) tested whether participation in a randomized controlled trial of the Parents as Teachers (PAT) psychosocial intervention (37) among families living in disadvantaged conditions was associated with reduced methylation at the NGFI-A binding domain (hereafter DNA methylation). The authors showed that PAT was associated with decreased DNA methylation, whereas parental disagreement (PD) and maternal depression symptoms were associated with increased DNA methylation and behavioral problems at age three. Furthermore, an indirect association between PD and internalizing symptoms via DNA methylation was observed.

The present study extends these findings in three key directions. First, we examine the prospective link between DNA methylation at age three and later cognitive outcomes at age five, which has not been previously investigated. Second, we test whether DNA methylation mediates the association between early stressors (parental disagreement) and cognitive performance. Third, we replicate the association between the PAT intervention and DNA methylation using an imputed dataset to handle missing data.

We report three main findings: (1) higher DNA methylation levels at age three are associated with lower cognitive performance at age five, providing preliminary empirical evidence for a prospective link between DNA methylation and cognitive outcomes in childhood; (2) DNA methylation mediates the association between parental disagreement and concentration problems; and (3) the PAT intervention is associated with lower DNA methylation levels. These findings may underscore the role of early psychosocial conditions and targeted home-visiting interventions in shaping epigenetic and cognitive development.

## Materials and Methods

This study is a secondary analysis of data from the Zurich Equity Prevention Project with Parents Participation and Integration (ZEPPELIN) RCT, which examines the impact of psychosocial and biological factors on development (38–40). This study employs a longitudinal observational design and includes two time points of data collection from ZEPPELIN RCT: T3, which includes DNA methylation analyses (as part of the ZEPPELIN 0-3 study, 2011–2015), and T5, which includes cognitive function assessments (as part of the ZEPPELIN 5-8 study, 2017–2020 (36)).

The ZEPPELIN study protocol was approved by the Ethics Committee of the canton of Zurich (Reference Nr. KEK-ZH 2013-0278). Written informed parental consent for all participating children was obtained between June 3, 2014, and July 8, 2015, in accordance with the principles of the Declaration of Helsinki. The trial was registered on ClinicalTrials.gov (NTC02882763).

### Participants

Participants in the current study are a subset drawn from the full ZEPPELIN cohort, as only families who signed informed consent to provide saliva samples of their child were included (see Fig. 1). The full ZEPPELIN cohort includes 248 psychosocially at-risk families identified through an interdisciplinary network of family counseling and health professionals in the metropolitan area of the city of Zurich at three project sites in the canton of Zurich. Families were recruited before or shortly after childbirth based on their answers on a risk-screening questionnaire (38). Risk factors were parental risks (low level of education, early parenthood, alcohol or drug abuse, sickness and disabilities), familial risks (single parenthood, partnership conflicts), social risks (lack of social integration, dissocial environment), material risks (unemployment, financial problems, confined living space) and child related risks (high-risk pregnancy, health issues, prematurity). Families were included if they showed at least two risk factors which were not compensated by protective factors, such as support from the extended family, stable and reliable parental figures, or clear and transparent family structures. Exclusion criteria were no permanent residency permit, severe illness or disability of the child, severe illness or disability of the parent requiring inpatient and long-term psychiatric treatment, and other intensive treatments or child protection procedures. After agreeing to participate in the study, families were assigned to the IG (n=132) or CG (n=116). The assignment was performed using stratified block randomization. Strata were project site, cumulative psychosocial risk factors assessed using the short screening, Swiss nationality, family structure (single parent: yes/no), and German-language proficiency (interpreter: yes/no) (39). From the 248 families recruited, 131 agreed to provide saliva samples of their children. A total of 135 saliva samples were collected (IG = 72, CG = 63). After biochemical analyses, three samples from the CG were excluded due to insufficient sequencing coverage. Among the excluded samples, one was a twin (from a twin pair) and two were from single-child families. Consequently, the final dataset includes N = 132 saliva samples (IG = 72, CG = 60).

**Fig. 1.**
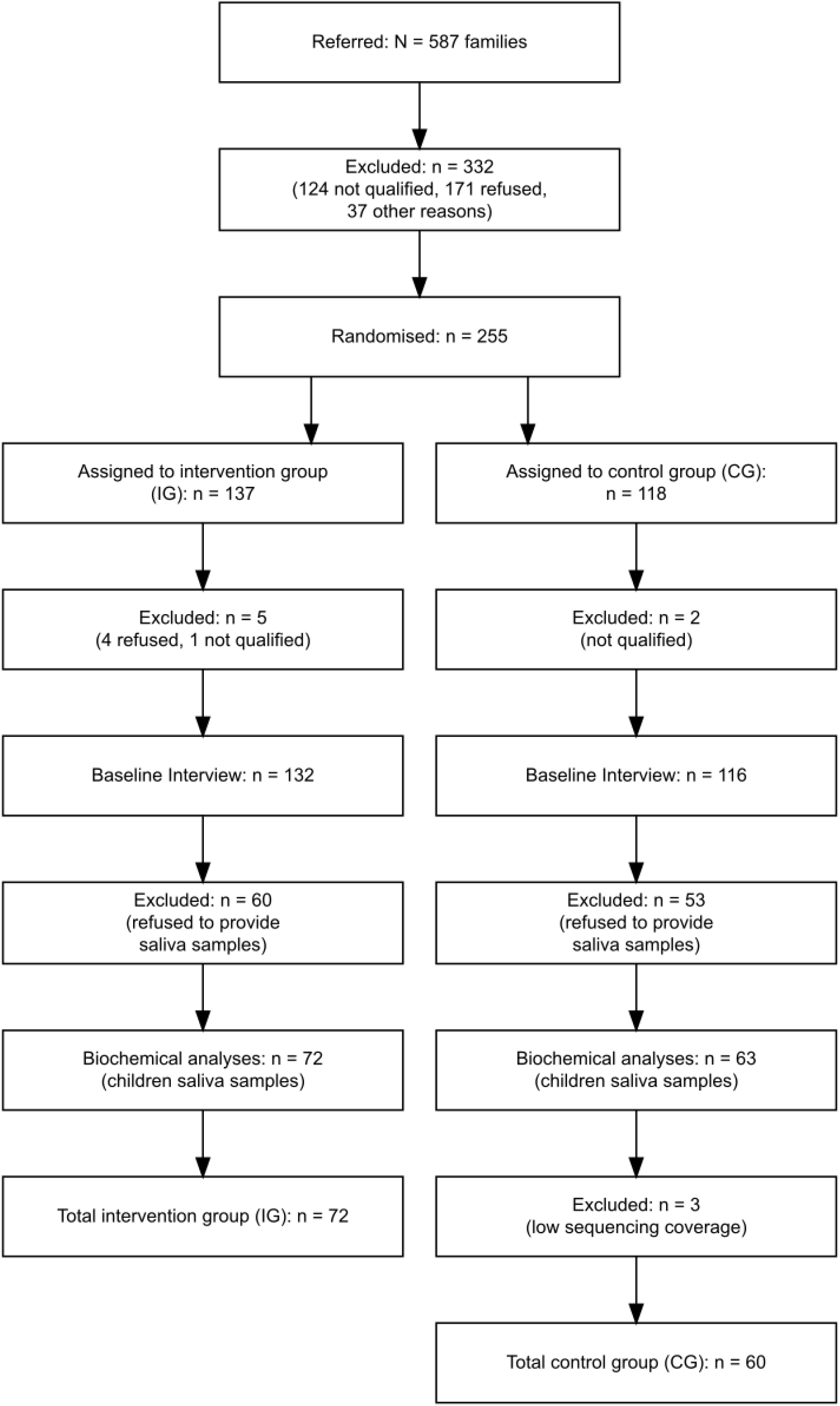
CONSORT flow diagram of participant recruitment and retention, modified from Schaub et al. (**39**)

At recruitment, all families lived in the Canton of Zurich, Switzerland, and 79.5% primarily spoke a non-German language (39).

### Parents as Teachers (PAT)

The PAT intervention is a home-based program that aims to improve parenting quality and provide enriched learning opportunities for children living in psychosocially disadvantaged conditions (41,42). By strengthening parental competencies, increasing knowledge of child development, and enhancing social support, this type of program also aims to reduce parental stress, decrease the risk of child abuse and neglect, and reduce early child behavior problems (37,43).

Participants in the IG received home visits from a trained parent educator throughout the child’s first three years, with at least ten visits per year. In the current sample, families participated in an average of n = 49.9 (SD = 10.1) home visits prior to data collection. Each visit focused on three key areas: fostering developmentally supportive parenting, enhancing parent–child interactions, and promoting overall family well-being. Additionally, parents were offered optional monthly group meetings to share experiences with other families and access information on parenting strategies, parent–child relationships, and community resources. The intervention also included monitoring of the child’s health and developmental progress, as well as guidance to support parents’ integration into the community and referrals to relevant services when needed. Families in the CG could use available community services but did not receive the PAT program. Both intervention and control families were offered incentives for participation, including birthday cards, small gifts, and a monetary compensation of 70 CHF for completing the study assessments.

### Data collection

#### Cognitive functions

The Snijders-Oomen Nonverbal (SON-R-2.5-7) Intelligence Test (44) was administered to assess cognitive functions in children at age five. This is a non-verbal intelligence test particularly suited for children with diverse language abilities. The test consists of activities that assess abstract reasoning (problem-solving, pattern recognition), visual-spatial skills (ability to manipulate and understand visual and spatial information), perceptual organization, memory and attention. Scores from each subset were combined to generate an overall IQ score (Cronbach’s α = 0.82).

As part of the standard SON-R procedure, examiners rated the child’s concentration, understanding of instructions, motivation, and cooperation on a 4-point ordinal scale (’good’, ‘variable’, ‘fair’, ‘poor’) immediately after test administration. The test manual recommends that examiners record these qualitative observations, as they can provide valuable context for interpreting the child’s performance (SON-R manual, p. 90). The examiners observed the following behaviors: (a) concentration – throughout the test, the examiner observed whether the child was attentive, easily distracted, needed to be called back to the task, and required many breaks; (b) understanding of instructions – the examiner assessed whether the child understood what to do after a non-verbal or verbal demonstration, started immediately or seemed confused, and asked for clarification; (c) motivation – the examiner noted whether the child was engaged and interested, tried to solve puzzles even when difficult, or gave up easily and lost interest; and (d) cooperation – the examiner considered whether the child followed instructions, accepted feedback ("No, not like that, look"), and tried to cooperate, or whether the child was oppositional or tried to run away.

All examiners (n = 12) were graduate students in early childhood education, applied psychology, or special education, and received specific training on the SON-R from the same instructor as part of the ZEPPELIN study. Training also included video feedback on test administration. The test was administered individually (1:1) and lasted 50 to 60 minutes.

No formal inter-rater reliability was assessed; however, the standardized training procedure ensured consistency across examiners. Following the manual’s recommendation that the validity of these observational ratings can be examined through their association with test performance (SON-R manual, p. 90), we assessed concurrent validity in our sample. All four observational ratings were significantly correlated with each other (rs = 0.32–0.58, p < .01) and with children’s IQ scores (rs = 0.27–0.41, p < 0.01), supporting their internal consistency and concurrent validity.

#### Early life stressors

Parental disagreement (PD), measured when the children were three years old, was used as an early life stressor based on its association with DNA methylation in our previous study (36) and its role in stress development (45–47). PD was measured through two items from the Parental Stress Questionnaire (PSQ) (48), assessing self-reported co-parenting disagreement on child upbringing. Specifically, the items “My partner and I totally agree on questions about upbringing” and “My partner and I discuss and decide together about parenting tactics” were rated by the mothers on a 4-point Likert scale (1 = strongly disagree to 4 = strongly agree), and a mean score was computed across the two items (Cronbach’s α = 0.81). To facilitate interpretation, scores were reversed for the statistical analyses so that higher scores indicated greater risk.

#### DNA methylation

Saliva was used to measure DNA methylation levels of children at age three. Salivary cells are easily obtainable and have been described as the peripheral tissues that best reflect brain-related processes (49). Saliva was collected in Oragene tubes (OG-500, DNA Genotek, Ontario, Canada) via the passive drool method, following the manufacturer’s guidelines, and stored at 4°C for later biochemical analysis. We used targeted bisulfite next-generation sequencing (NGS) to assess methylation at all five CpGs within the NGFI-A binding domain, which was not feasible with genome-wide BeadChip arrays, as only one of the five CpGs of interest was included in the array. Genomic DNA was isolated using the PrepIT-L2P protocol (OG-500, DNA Genotek, Ontario, Canada). DNA bisulfite conversion was achieved using the EZ DNA methylation kit (Zymo Research, Irvine, CA, USA) following the manufacturer’s instructions. Primers used for the amplification of the targeted sequence (Fig. 2, reprinted from Gardini et al. (36)) were as follows: forward primer 5′-TTG AAG TTT TTT TAG AGG G-3′ and reverse primer 5′-AAT TTC TCC AAT TTC TTT TCT C-3′. Libraries for Illumina sequencing were prepared using the v3 kit (Illumina, San Diego, CA, USA) and sequenced at the Genetic Diversity Centre of the ETH, Zurich, Switzerland. The software Trimmomatic v0.35 (licensed under GPL V3) was employed to identify and remove low-quality products according to Bolger et al. (50). The mean coverage for all the analyzed CpGs was around 50,000X. The Bismark program (v0.19.0) was used to extract the counts of methylated (cytosines) and unmethylated (thymine) bases. For each CpG position, a percentage of methylation was calculated. To assess the overall methylation status of the NGFI-A binding region (hereafter referred to as DNA methylation), the mean methylation level across the five CpG sites within this transcription factor binding domain was computed.

**Fig. 2.**
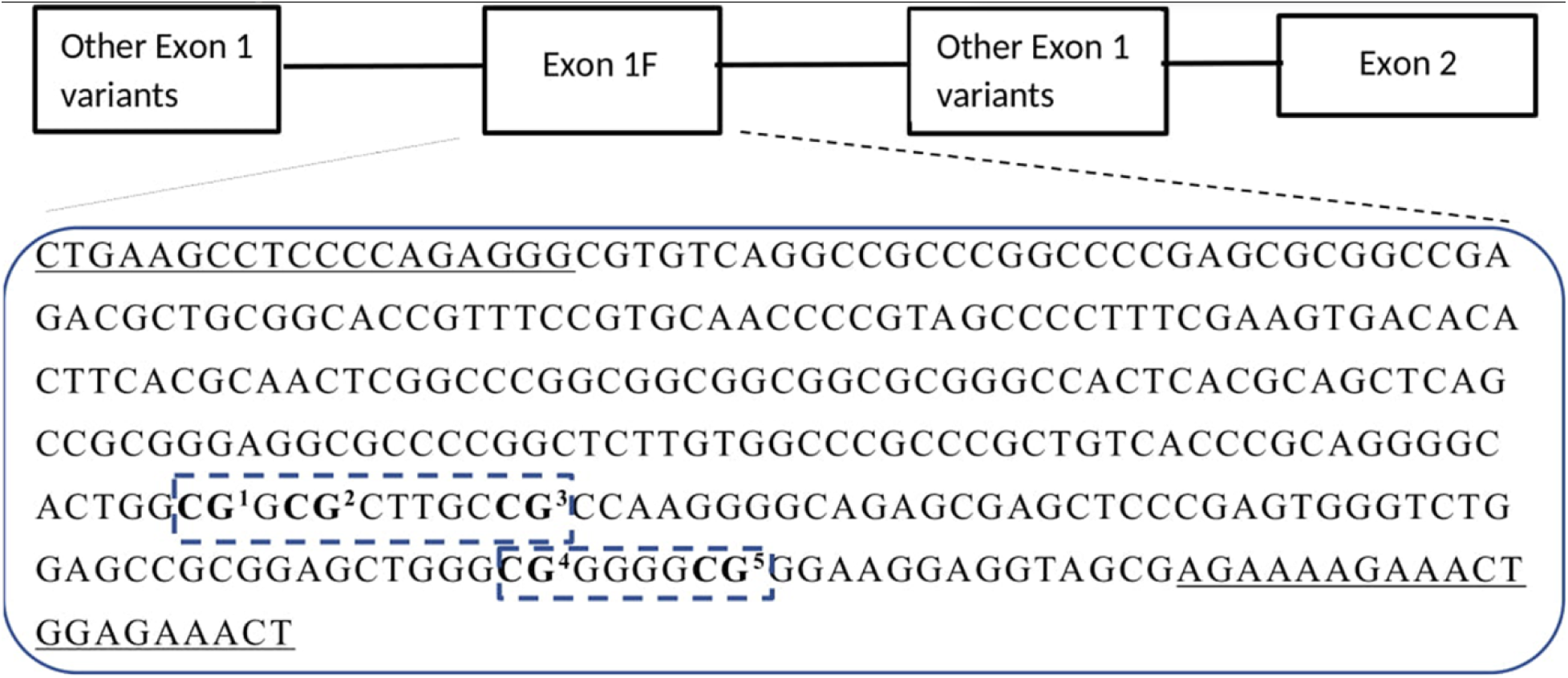
**Depiction of the *NR3C1* exon 1F sequence,** located at chr5 (hg38):143,404,021–143,404,338 within the 5′ untranslated region of the *NR3C1* gene. The NGFI-A binding region is highlighted with dashed boxes. This figure is reprinted from Gardini et al. (36).

### Statistical analyses

Descriptive statistics, including counts, percentages, means, and standard deviations, were computed for all study variables. Pearson correlations were conducted to examine bivariate associations between variables.

Linear regression analyses were performed to test the effect of DNA methylation on cognitive functions and the intervention on DNA methylation. To examine whether the direct association between methylation and IQ followed a nonlinear or threshold-like pattern, we tested quadratic, spline, and segmented regression models. Regression-based mediation analyses using the mediation package in R (with 1,000 bootstrap resamples) were conducted to examine the pathways linking the intervention, parental disagreement, DNA methylation, and cognitive functions. All mediators were included in the mediation analyses if they showed significant associations with both the independent and dependent variables. Following the recommendations of Hayes (51) and Zhao et al. (52), we adopted an indirect-only mediation approach and tested indirect effects regardless of the significance of the direct effects. In this framework, mediation is established solely by the significance of the indirect effect, and a significant direct effect is not required as a precondition. Zhao et al. (52) explicitly classify ‘indirect-only mediation’ as a valid pattern in which the indirect effect is significant while the direct effect is not.

Covariates included standard variables—age, gender, and region of origin—as well as socio-economic status (SES) as measured by ISEI (53) and birth weight, due to their previously reported associations with cognitive functions and DNA methylation (17,54–56). Because IQ scores were already standardized for age, age at T5 was included as a covariate only in models examining observational outcomes as dependent variables (e.g., concentration, understanding of instructions), not in models with IQ as the dependent variable. Models examining the effect of the intervention were adjusted for child’s age at randomization and family outreach difficulty, due to their significant association with the group variable in this sample (Table 2). Family outreach difficulty has been described as a major indicator of greater family need (38) and in this study sample was significantly more common in the CG.

DNA methylation values were log-transformed to improve normality due to a strong positive skew. For all continuous and ordinal variables, z-scores were computed. To handle missing data, we employed multiple imputation (MI) under the Missing At Random (MAR) assumption using Predictive Mean Matching (PMM). A total of 20 imputed datasets were generated, and results were combined using Rubin’s rules. Across all variables, 6.5% of data points were missing. Missingness was concentrated in parental disagreement (16.7%), cognitive functions (15.2%) and SES (12.1%). Other variables had minimal (<3%) or no missing data. To assess the robustness of our results, we conducted complete-case analysis (listwise deletion) and Missing Not At Random (MNAR) analyses applying a delta adjustment of +1 standard deviation to imputed values of parental disagreement (higher values indicating more disagreement) and –1 standard deviation to imputed values of socioeconomic status (lower values indicating lower SES). Statistical significance was set at α = 0.05 (two-tailed). All statistical analyses were performed using R, version 4.3.1.

## Results

### Descriptive statistics

The analytical sample of the present study (*N* = 132) did not differ from the full ZEPPELIN cohort (*N* = 248) regarding the strata used for the randomization process, nor regarding child age at recruitment, sex, and SES (see supporting information).

Descriptive statistics (counts, percentages, means, and standard deviations) for all study variables are presented in Table 1. The mean age at randomization (T0) was 50.9 days (*SD* = 40.7), the mean age at saliva sampling (T3) was 35.5 months (*SD* = 1.1), and the mean age of IQ assessment (T5) was 65.6 months (*SD* = 3.9). Most children were born with a birth weight of ≥2.5 kg (86.3%), had European origins (71.2%), and the family mean SES was 34.4 (SD=23.1), indicating a middle-to-low socioeconomic level. The mean IQ score was 94.8 (*SD* = 14.7), 19 children scored below 80 (5 below 70), while 4 children scored above 120. Ratings of problems in concentration, understanding of instructions, motivation, and cooperation were generally low, with concentration showing the greatest variability. Mean PD was 3.3 (*SD* = 0.7), indicating moderate parental disagreement. Mean DNA methylation was 0.5 (SD = 1.7). Pearson correlations among all study variables are presented in Table 2. Among significant results, higher IQ was negatively correlated with concentration, understanding, motivation, and cooperation difficulties as expected, and positively correlated with SES and birth weight. DNA methylation showed positive correlations with concentration difficulties and PD.

**Table 1.** Descriptive statistics.

| **Sociodemographic and perinatal variables** |  |
| --- | --- |
| PAT Group, n (%) |  |
| 1= Intervention group (IG) | 72 (55%) |
| 0= Control group (CG) | 60 (45%) |
| Gender, n (%) |  |
| 1= Girls | 72 (55%) |
| 0= Boys | 60 (45%) |
| Age at T0 (days), Mean ± SD | 50.9 ± 40.7 |
| Birth weight (g), Mean ± SD | 3200 ± 621 |
| Geographical Origin, n (%) |  |
| 1= Europe | 94 (71.2%) |
| 0= Other (Africa, Asia, Latin America) | 38 (29.8%) |
| SES, Mean ± SD | 34.4 ± 23.1 |
| **Biological assessment** |  |
| DNA methylation, Mean ± SD | 0.5 ± 1.7 |
| Age at T3 (months), Mean ± SD | 35.5 ± 1.1 |
| **Cognitive assessments** |  |
| IQ, Mean ± SD | 94.8 ± 14.7 |
| **Observational assessments (1 = good to 4 = poor)** |  |
| Concentration, Mean ± SD | 1.8 ± 0.8 |
| Understanding of the instructions, Mean ± SD | 1.2 ± 0.5 |
| Motivation, Mean ± SD | 1.5 ± 0.7 |
| Cooperation, Mean ± SD | 1.3 ± 0.6 |
| Age at T5 (months), Mean ± SD | 65.6 ± 3.9 |
| **Parental and family variables** |  |
| Parental disagreement, Mean ± SD | 3.3 ± 0.7 |
| Family outreach difficulty, n (%) |  |
| Yes=0 | 16 (12.1%) |
| No=1 | 116 (87.9%) |

**Table 2.** Bivariate Pearson correlation between all study variables.

| Variable | M | SD | PAT Group | Gender | Age T0 | Birth weight | Origin | SES | Methylation | Age T3 | IQ | Concentration prob. | Understanding prob. | Motivation prob. | Cooperation prob. | Age T5 |
| --- | --- | --- | --- | --- | --- | --- | --- | --- | --- | --- | --- | --- | --- | --- | --- | --- |
| PAT Group | 0.55 | 0.50 |  |  |  |  |  |  |  |  |  |  |  |  |  |  |
| Gender | 0.55 | 0.50 | 0.14 |  |  |  |  |  |  |  |  |  |  |  |  |  |
| Age T0 | 50.89 | 40.74 | -0.09 | -0.08 |  |  |  |  |  |  |  |  |  |  |  |  |
| Birth weight | 3,189.80 | 630.98 | -0.10 | -0.09 | 0.17* |  |  |  |  |  |  |  |  |  |  |  |
| Origin | 0.71 | 0.45 | 0.09 | 0.02 | 0.06 | -0.17* |  |  |  |  |  |  |  |  |  |  |
| SES | 33.97 | 23.26 | -0.07 | 0.00 | 0.20* | -0.02 | 0.28** |  |  |  |  |  |  |  |  |  |
| Methylation | -0.82 | 0.48 | -0.13 | 0.05 | 0.21* | -0.00 | 0.05 | 0.04 |  |  |  |  |  |  |  |  |
| Age T3 | 1,101.21 | 33.59 | -0.18* | -0.03 | -0.03 | 0.06 | -0.24** | -0.14 | -0.13 |  |  |  |  |  |  |  |
| IQ | 94.67 | 14.46 | 0.03 | 0.17 | -0.09 | 0.18* | -0.03 | 0.27** | -0.09 | 0.05 |  |  |  |  |  |  |
| Concentration prob. | 1.68 | 0.78 | -0.08 | -0.20* | 0.11 | 0.00 | -0.00 | -0.10 | 0.26** | -0.11 | -0.39*** |  |  |  |  |  |
| Understanding prob. | 1.16 | 0.48 | 0.02 | -0.01 | -0.03 | -0.10 | -0.03 | -0.18* | 0.16 | -0.03 | -0.37*** | 0.28** |  |  |  |  |
| Motivation prob. | 1.42 | 0.71 | -0.24** | -0.06 | -0.00 | -0.01 | -0.12 | -0.21* | 0.07 | -0.07 | -0.43*** | 0.54*** | 0.21* |  |  |  |
| Cooperation prob. | 1.24 | 0.59 | -0.19* | -0.09 | 0.11 | 0.03 | -0.16 | -0.23** | -0.01 | 0.26** | -0.27** | 0.49*** | 0.24** | 0.52*** |  |  |
| Age T5 | 65.71 | 3.91 | -0.12 | -0.09 | 0.33*** | -0.17 | -0.12 | 0.02 | -0.01 | 0.15 | -0.13 | 0.01 | 0.08 | -0.04 | 0.10 |  |
| PD | 3.32 | 0.70 | 0.05 | -0.10 | 0.09 | -0.02 | -0.15 | -0.22* | -0.29*** | 0.02 | -0.02 | -0.08 | -0.04 | -0.07 | 0.00 | 0.16 |

Note: \* p < 0.05, \*\* p < 0.01, \*\*\* p < 0.001. M = mean; SD = standard deviation.

### Linear regressions

We found no significant association between methylation levels and IQ scores (*β*=−0.083, *t*(112)=−0.974, p*=*0.332). No nonlinear or threshold effects were detected, and neither spline nor segmented regression improved model fit. However, higher methylation levels were associated with difficulties in concentrating (*β*=0.244, *t*(117)=2.849, p=0.005). In addition, higher IQ scores were associated with higher SES (*β*=0.230, *t*(81)=2.399, p=0.019) and birth weight (*β*=0.191, *t*(96)=2.106, p=0.038), while being a boy was associated with higher concentration difficulties (*β*=-0.438, *t*(86)=-2.315, p*=*0.023). No statistically significant associations were observed in models including understanding of the instructions, cooperation and motivation as dependent variables.

The intervention was associated with lower methylation levels (*β*=-0.375, *t*(121)=-2.068, p=0.041). In addition, starting the intervention later was significantly associated with higher levels of DNA methylation (*β*=0.465, *t*(121)=2.640, p=0.009).

### Mediation analyses

The indirect effect of the intervention group on concentration via methylation was not statistically significant (indirect effect = −0.085, p = 0.087), suggesting no mediation. DNA methylation levels were indirectly associated with IQ scores through concentration problems (indirect effect = −0.083, p = 0.012; direct effect = 0.004, p = 0.802; total effect = −0.079, p = 0.802). In addition, PD was significantly associated with concentration difficulties through methylation (indirect effect = 0.064, p = 0.023; direct effect = 0.051, p = 0.602; total effect = 0.115, p = 0.602). The indirect effect did not significantly differ between the intervention and control groups (difference = −0.016, p = 0.531), indicating no moderated mediation.

Finally, the serial indirect effect of parental disagreement on IQ through increased DNA methylation and increased concentration problems was not significant (β = −0.024, p = 0.119).

Results were largely consistent across multiple imputation (MAR) and complete-case analyses, with two exceptions. First, the effect of the intervention on methylation was significant under MAR (β = −0.375, p = 0.041) but not in complete cases (β = 0.015, p = 0.949). Second, the indirect effect of parental disagreement on concentration via methylation was significant under MAR (indirect effect = 0.064, p = 0.023) but only marginal in complete cases (indirect effect = 0.076, p = 0.076). Regarding covariates, socioeconomic status and birth weight were significantly associated with IQ in both MAR and complete-case analyses (SES: β = 0.23-0.26, p < 0.05; birth weight: β = 0.19-0.23, p < 0.05). Gender was significantly associated with concentration in both models (β = −0.44 to −0.46, p < 0.05), with girls showing fewer concentration difficulties. Child’s age at randomization was consistently associated with higher methylation levels (β = 0.24-0.29, p < 0.01), indicating that later study entry (i.e., older age at randomization) was related to increased DNA methylation. However, this effect did not differ between the intervention and control groups (interaction β = 0.055, p = 0.756).

Under the MNAR assumption (imputing missing SES values one standard deviation below the observed mean and missing parental disagreement values one standard deviation above the observed mean), results remained largely unchanged. Specifically, the effects of the intervention on methylation (β = −0.367, p = 0.043), the indirect effect of methylation on IQ via concentration (indirect effect = −0.078, p = 0.032), and the indirect effect of parental disagreement on concentration via methylation (indirect effect = 0.054, p = 0.032) remained significant. The association between methylation and understanding of instructions became significant under MNAR (β = 0.183, p = 0.037), while it was marginal under MAR (p = 0.062). All other conclusions, including the absence of moderated mediation and non-significant serial mediation, were consistent across both missing data assumptions.

## Discussion

Family-level psychosocial adversity is a key predictor of children’s later socioeconomic disparities, often emerging early through differences in academic achievement (1,2). This RCT examined DNA methylation at the NGFI-A binding site of the *NR3C1* 1F promoter as a potential biological mechanism contributing to these disparities.

The main finding was that DNA methylation at age three was associated with greater concentration problems at age five, which in turn were associated with lower IQ. An indirect association between methylation and IQ via concentration problems was observed, while the direct association was not significant. This pattern indicates that the observed association between methylation and IQ operates entirely through concentration difficulties (indirect-only mediation; (52)). In line with previous findings, these results may suggest that an early increase in methylation at the NGFI-A binding domain of the *NR3C1* 1F promoter is associated with a maladaptive regulation of the HPA axis that is maintained over time, potentially undermining the capacity to concentrate in the long term or eliciting situational-dependent abnormal stress responses (e.g., test anxiety) that interfere with attention (8,10,11,28–34). These mechanisms are likely to be mediated by prefrontal functioning (57).

Second, in our study, PD was associated with concentration problems through DNA methylation. Previously, negative co-parenting interactions have been shown to lead to high levels of parental stress, which has been associated with children’s psychological maladjustment, impaired cognitive development, sleep problems and higher average cortisol levels, contributing to fatigue, illness, and sleep disturbances (58–61). Our results add to these findings by suggesting that DNA methylation may be involved as an underlying mechanism of these associations, specifically linking PD to concentration problems. In this study, concentration problems were assessed through direct observation of the child’s behavior. If replicated, this finding could have practical implications for caregivers and educators. For instance, attention difficulties that might otherwise be mistaken for laziness or low academic potential could, in some cases, reflect physiological adaptations of the stress system. Recognizing these signs early might, in principle, allow for timely interventions to prevent potential long-term consequences. However, these interpretations remain highly speculative and require replication in independent samples using validated measures.

Third, our study also provides further evidence for the association between the PAT intervention and DNA methylation, which was already shown in Gardini et al. (36) using model-specific complete-case analysis. Participation in the three-year program was associated with lower DNA methylation levels at the end of the intervention. Previous research has shown that PAT positively affects child development, in part by promoting a less stressful family environment (37,43). Our findings suggest that epigenetic modifications in genes regulating the HPA axis response may represent one possible biological mechanism underlying these effects. However, although the intervention was associated with reduced DNA methylation, the indirect pathway from the intervention to cognitive functions via methylation only reached marginal statistical significance. It is possible that the magnitude of the biological change was insufficient to translate into measurable differences in cognitive outcomes within the current sample size, or that larger samples are needed to reliably detect this effect. In addition, the significant association between the child’s age at randomization (i.e., time of entry into the study) and DNA methylation levels at age three may indicate that earlier study entry is associated with lower methylation levels. This finding would be consistent with previous evidence suggesting that interventions should be implemented as early as possible to counteract maladaptive epigenetic changes in early life that may contribute to the long-term effects of early stress (1). However, the absence of a significant interaction between group assignment and age at randomization may also suggest that non-specific aspects of study participation (e.g., being enrolled in a longitudinal study, such as increased parental awareness, regular monitoring, or perceived support from the research team) may have influenced DNA methylation.

Our results also confirmed previous findings: higher birth weight and higher SES were associated with higher IQ, whereas boys exhibited more concentration problems compared to girls (17,54,62).

To the best of our knowledge, this is the first study to report an association between early DNA methylation at the NGFI-A binding site of the *NR3C1* 1F promoter and later cognitive functions in children. These findings highlight the potential role of biological stress regulation as a mechanism through which early environmental conditions may influence cognitive development. However, given the exploratory nature of this study and the study limitations, these findings should be interpreted with caution. Future research should aim to replicate these results in larger, independent samples and investigate whether early interventions can be optimized to more effectively prevent maladaptive epigenetic programming.

### Limitations

A primary limitations of the study is the absence of DNA methylation assessment at T5. This would have allowed for a better understanding of the relative contribution of early versus later methylation patterns to cognitive functions, as well as the stability of early DNA methylation changes over time. Another limitation concerns the relatively small sample size, which may have reduced the power to detect small indirect or moderated effects, particularly in multi-path models. The assessment of concentration problems, understanding of instructions, motivation, and cooperation was based on examiner ratings following the SON-R test battery. While this is a standard procedure in the SON-R, these ratings have several inherent limitations. The ratings are subjective, as they rely on the examiner’s judgment rather than on objective behavioral metrics. The ratings reflect a single time point (immediately after test administration), which may not capture variability in a child’s attention across different contexts or over time. Situational factors such as the child’s fatigue, mood, or test anxiety on the day of assessment could have influenced the ratings. Given the limitations outlined above, findings involving concentration problems should be interpreted with caution. Future studies should adopt a multi-method approach to determine whether DNA methylation may serve as a reliable biomarker for concentration problems, integrating repeated assessments of both methylation and attention, observational ratings across multiple time points, parent- or teacher-report questionnaires, and—where feasible—performance-based attention measures to better capture the stability and ecological validity of the observed associations. Finally, physiological markers of HPA axis activity (e.g., cortisol levels) were not included, which limits the possibility of directly linking methylation patterns with functional stress responses.

## Acknowledgments

The data reported and analyzed in this paper were generated in collaboration with the Genetic Diversity Centre (GDC), ETH Zurich

