## Supplementary information for "Early DNA methylation at the NGFI-A binding site of the *NR3C1* 1F promoter predicts cognitive functions at age five: evidence from the Parents as Teachers intervention in the ZEPPELIN study"

Study variables

| | |
|---|---|
| **cngrup** | PAT Group |
| **cnsex** | Gender |
| **cnrandalt** | Age at T0 |
| **cngew** | Birth weight |
| **Europe_vs_Others** | Geographical Origin |
| **h3isei** | Socio-economic status (SES) |
| **logMeth** | DNA methylation |
| **h3tage** | Age at T3 |
| **son5iq** | IQ |
| **son5kon** | Concentration |
| **son5ver** | Understanding of the instructions |
| **son5mot** | Motivation |
| **son5koo** | Cooperation |
| **son5mon** | Age at T5 |
| **e3part** | Parental disagreement |
| **cnerr** | Family outreach difficulty |

### Analytical study sample vs full cohort comparison

#### Sex, SES, age at randomization

```
p_sex <- chisq.test(table(dati_zeppelin$group, dati_zeppelin$cnsex))$p.value
> p_hisei <- wilcox.test(hisei ~ group, data = dati_zeppelin)$p.value
> p_rand <- wilcox.test(cnrandalt ~ group, data = dati_zeppelin)$p.value
> results <- data.frame(
+   Variable = c("cnsex", "hisei", "cnrandalt"),
+   Type = c("Categorical", "Continuous", "Continuous"),
+   Test = c("Chi-square", "Wilcoxon", "Wilcoxon"),
+   p_value = c(p_sex, p_hisei, p_rand)
+ )
> results$Interpretation <- ifelse(results$p_value < 0.05,
+                                  "SIGNIFICANT difference",
+                                  "NOT significant (comparable)")
> print(results)
   Variable        Type       Test   p_value
1     cnsex Categorical Chi-square 0.6720438
2     hisei  Continuous   Wilcoxon 0.5827588
```

```
3 cnrandalt  Continuous   Wilcoxon 0.4456485
                Interpretation
1 NOT significant (comparable)
2 NOT significant (comparable)
3 NOT significant (comparable)
```

**Project site, german proficiency (interpreter YES/NO), family structure (single parent), stress-HBS**

```
# Extract p-values
> p_cnsort <- chisq.test(table(dati_zeppelin$group, dati_zeppelin$cnsort))$p.
value
> p_cnscre1 <- chisq.test(table(dati_zeppelin$group, dati_zeppelin$cnscre1))$
p.value
> p_cnscre4b <- chisq.test(table(dati_zeppelin$group, dati_zeppelin$cnscre4b)
)$p.value
> # For continuous variable hbelges (using Wilcoxon as default)
> p_hbelges <- wilcox.test(hbelges ~ group, data = dati_zeppelin)$p.value
> # Create results table
> results <- data.frame(
+   Variable = c("cnsort", "cnscre1", "cnscre4b", "hbelges"),
+   Type = c("Categorical", "Dichotomous", "Dichotomous", "Continuous"),
+   Test = c("Chi-square", "Chi-square", "Chi-square", "Wilcoxon"),
+   p_value = c(p_cnsort, p_cnscre1, p_cnscre4b, p_hbelges)
+ )
> # Interpretation
> results$Interpretation <- ifelse(results$p_value < 0.05,
+                                   "SIGNIFICANT difference",
+                                   "NOT significant (comparable)")
> print(results)
  Variable        Type       Test   p_value
1   cnsort Categorical Chi-square 0.2145486
2  cnscre1 Dichotomous Chi-square 0.9124248
3 cnscre4b Dichotomous Chi-square 1.0000000
4  hbelges  Continuous   Wilcoxon 0.7368180
                Interpretation
1 NOT significant (comparable)
2 NOT significant (comparable)
3 NOT significant (comparable)
4 NOT significant (comparable)
```

**cnrand (Types of randomization by stratification characteristics)**

```
> print(chisq_cnrand)

        Pearson's Chi-squared test

data:  tab_cnrand
X-squared = 1.4949, df = 6, p-value = 0.9598

> # Fisher's exact test if expected counts < 5
```

```
> fisher.test(tab_cnrand)

        Fisher's Exact Test for Count Data

data:  tab_cnrand
p-value = 0.9629
alternative hypothesis: two.sided
```

```
> print(results)
  Variable        Type       Test   p_value
1   cnsort Categorical Chi-square 0.2145486
2  cnscre1 Dichotomous Chi-square 0.9124248
3 cnscre4b Dichotomous Chi-square 1.0000000
4   cnrand Categorical Chi-square 0.9598333
5  hbelges  Continuous   Wilcoxon 0.7368180
              Interpretation
1 NOT significant (comparable)
2 NOT significant (comparable)
3 NOT significant (comparable)
4 NOT significant (comparable)
5 NOT significant (comparable)
```

**MICE imputation under MAR assumption**

```
# ============================================================
> # MICE IMPUTATION
> # ============================================================
>
> # Step 1: Load libraries and raw data
> library(mice)
> library(dplyr)
>
> # Load raw data (with dummy codes -444, -555, etc.)
> raw_data <- read.csv("original dataset.csv")
>
> # ============================================================
> # Step 2: Convert dummy codes to NA
> # ============================================================
>
> dummy_codes <- c(-333, -444, -555, -777, -999)
>
> data_clean <- raw_data
>
> for(code in dummy_codes) {
+   data_clean[data_clean == code] <- NA
+ }
>
> # Check missing counts
> cat("Missing values after conversion:\n")
Missing values after conversion:
> missing_count <- colSums(is.na(data_clean))
```

```
> print(missing_count[missing_count > 0])
 h3tage  h3isei   cngew  e3part  son5iq son5kon son5ver son5mot son5koo son5mon 
      3      16       1      22      20      20      20      20      20      20 
> 
> cat("\nTotal N:", nrow(data_clean), "\n")

Total N: 132 
> 
> # ================================================================
> # Step 2b: Calculate exact missing percentages
> # ================================================================
> 
> missing_percent <- colSums(is.na(data_clean)) / nrow(data_clean) * 100
> missing_percent_sorted <- sort(missing_percent[missing_percent > 0], decreasing = TRUE)
> 
> cat("=== MISSING PERCENTAGES ===\n")
=== MISSING PERCENTAGES ===
> print(round(missing_percent_sorted, 1))
 e3part  son5iq son5kon son5ver son5mot son5koo son5mon  h3isei  h3tage   cngew 
   16.7    15.2    15.2    15.2    15.2    15.2    15.2    12.1     2.3     0.8 
> 
> # Key variables
> key_vars <- c("e3part", "son5iq", "son5kon", "son5ver", "son5mot", "son5koo", 
"son5mon", "h3isei")
> 
> for(var in key_vars) {
+   if(var %in% names(missing_percent_sorted)) {
+     cat(var, ": ", round(missing_percent_sorted[var], 1), "%\n", sep = "")
+   }
+ }
e3part: 16.7%
son5iq: 15.2%
son5kon: 15.2%
son5ver: 15.2%
son5mot: 15.2%
son5koo: 15.2%
son5mon: 15.2%
h3isei: 12.1%
> 
> # Overall missingness
> total_cells <- nrow(data_clean) * ncol(data_clean)
> missing_cells <- sum(is.na(data_clean))
> overall_missing_pct <- missing_cells / total_cells * 100
> 
> cat("\n=== OVERALL MISSINGNESS ===\n")

=== OVERALL MISSINGNESS ===
> cat("Total missing cells:", missing_cells, "out of", total_cells, "\n")
Total missing cells: 162 out of 2508 
> cat("Overall missing percentage:", round(overall_missing_pct, 1), "%\n")
Overall missing percentage: 6.5 %
> 
> # ================================================================
```

```
> # Step 3: Select variables for imputation
> # ============================================================
> 
> data_impute <- data_clean %>%
+   dplyr::select(cngrup, cnsex, h3tage, cnrandalt, h3isei, cngew,
+                 Europe_vs_Others, cnerr, logMeth, e3part,
+                 son5iq, son5kon, son5ver, son5mot, son5koo, son5mon)
> 
> cat("\nVariables selected for imputation:\n")

Variables selected for imputation:
> print(names(data_impute))
 [1] "cngrup"           "cnsex"            "h3tage"           "cnrandalt"       
"h3isei"           "cngew"           
 [7] "Europe_vs_Others" "cnerr"            "logMeth"          "e3part"          
"son5iq"           "son5kon"         
[13] "son5ver"          "son5mot"          "son5koo"          "son5mon"         
> 
> # ============================================================
> # Step 4: Multiple Imputation with PMM (20 imputations)
> # ============================================================
> 
> set.seed(2026)
> 
> imp <- mice(data_impute,
+             method = "pmm",
+             m = 20,
+             maxit = 20,
+             printFlag = TRUE)


> 
> # Save imputation object
> saveRDS(imp, file = "imputation_20_PMM_h3isei.rds")
> 
> cat("\n=== IMPUTATION COMPLETE ===\n")

=== IMPUTATION COMPLETE ===
> cat("Number of imputations:", imp$m, "\n")
Number of imputations: 20
> cat("Number of iterations:", imp$maxit, "\n")
Number of iterations: 
> 
> # ============================================================
> # Step 5: Create z-scores on imputed data
> # ============================================================
> 
> imp_long <- complete(imp, action = "long", include = TRUE)
> 
> add_z_scores <- function(data) {
+ 
+   data$z_h3tage <- as.numeric(scale(data$h3tage))
+   data$z_h3isei <- as.numeric(scale(data$h3isei))  # <--- CHANGED: was z_hisei
```

```
+   data$z_logMeth <- as.numeric(scale(data$logMeth))
+   data$z_e3part <- as.numeric(scale(data$e3part))
+   data$z_son5iq <- as.numeric(scale(data$son5iq))
+   data$z_son5kon <- as.numeric(scale(data$son5kon))
+   data$z_son5ver <- as.numeric(scale(data$son5ver))
+   data$z_son5mot <- as.numeric(scale(data$son5mot))
+   data$z_son5koo <- as.numeric(scale(data$son5koo))
+   data$z_son5mon <- as.numeric(scale(data$son5mon))
+   data$z_cngew <- as.numeric(scale(data$cngew))
+   data$z_cnrandalt <- as.numeric(scale(data$cnrandalt))
+
+   return(data)
+ }
>
> imp_transformed <- imp_long %>%
+   group_by(.imp) %>%
+   group_modify(~ as.data.frame(add_z_scores(.x))) %>%
+   ungroup()
>
> imp_mids <- as.mids(imp_transformed)
>
> # Save transformed object
> saveRDS(imp_mids, file = "imputation_20_PMM_h3isei_transformed.rds")
>
> cat("\n=== TRANSFORMATION COMPLETE ===\n")

=== TRANSFORMATION COMPLETE ===
> cat("Z-scores added for continuous/ordinal variables including z_h3isei\n")
Z-scores added for continuous/ordinal variables including z_h3isei
> cat("Categorical variables unchanged: cngrup, cnsex, Europe_vs_Others, cnerr\n")
Categorical variables unchanged: cngrup, cnsex, Europe_vs_Others, cnerr
>
> REUSLTS
>
> # ============================================================
> # MODELS 1-4: Linear Regressions
> # ============================================================
>
> library(mice)
> library(dplyr)
>
> imp <- readRDS("imputation_20_PMM_h3isei_transformed.rds")
>
> # ============================================================
> # MODEL 1: z_logMeth -> z_son5iq
> # Covariates: cnsex, Europe_vs_Others, z_h3isei, z_cngew
> # ============================================================
>
> model1 <- with(imp, lm(z_son5iq ~ z_logMeth + cnsex + Europe_vs_Others + z_h3isei +
z_cngew))
> pooled_model1 <- pool(model1)
> cat("\n========== MODEL 1 ==========\n")
```

```
========== MODEL 1 ==========
> print(summary(pooled_model1, conf.int = FALSE))
             term    estimate  std.error  statistic        df    p.value
1     (Intercept) -0.06171207 0.19706178 -0.3131610  95.84985 0.75483874
2       z_logMeth -0.08373626 0.08600009 -0.9736764 111.57934 0.33232338
3           cnsex  0.34385157 0.17958014  1.9147527  96.17506 0.05849588
4 Europe_vs_Others -0.16396301 0.21338892 -0.7683764  83.56880 0.44442954
5        z_h3isei  0.23048415 0.09607416  2.3990233  81.64784 0.01871701
6         z_cngew  0.19165870 0.09099575  2.1062378  96.45630 0.03777987
>
> # ============================================================
> # MODEL 2: z_logMeth -> z_son5kon
> # Covariates: cnsex, Europe_vs_Others, z_h3isei, z_cngew, z_son5mon
> # ============================================================
>
> model2 <- with(imp, lm(z_son5kon ~ z_logMeth + cnsex + Europe_vs_Others + z_h3isei +
z_cngew + z_son5mon))
> pooled_model2 <- pool(model2)
> cat("\n========== MODEL 2 ==========\n")

========== MODEL 2 ==========
> summary_df2 <- summary(pooled_model2, conf.int = FALSE)
> summary_df2$df <- round(summary_df2$df, 0)
> print(summary_df2)
             term     estimate  std.error   statistic  df     p.value
1     (Intercept)  0.202468530 0.19925532  1.01612607 104 0.311937278
2       z_logMeth  0.244052666 0.08564894  2.84945328 117 0.005174793
3           cnsex -0.438023739 0.18917770 -2.31540895  86 0.022967366
4 Europe_vs_Others  0.026495575 0.21268153  0.12457864  94 0.901124499
5        z_h3isei -0.108314688 0.09999338 -1.08321855  73 0.282263366
6         z_cngew  0.009856486 0.09216494  0.10694398 104 0.915039152
7       z_son5mon -0.005489188 0.09540601 -0.05753503  91 0.954244678
>
> # ============================================================
> # MODEL 3: z_logMeth -> z_son5ver, z_son5mot, z_son5koo
> # Covariates: cnsex, Europe_vs_Others, z_h3isei, z_cngew, z_son5mon
> # ============================================================
>
> # z_son5ver
> model3_ver <- with(imp, lm(z_son5ver ~ z_logMeth + cnsex + Europe_vs_Others + z_h3isei
> pooled_model3_ver <- pool(model3_ver)
> cat("\n========== MODEL 3: z_logMeth -> z_son5ver ==========\n")

========== MODEL 3: z_logMeth -> z_son5ver ==========
> summary_df3_ver <- summary(pooled_model3_ver, conf.int = FALSE)
> summary_df3_ver$df <- round(summary_df3_ver$df, 0)
> print(summary_df3_ver)
             term    estimate  std.error  statistic  df    p.value
1     (Intercept)  0.09615772 0.21893301  0.4392107  81 0.66168391
2       z_logMeth  0.17078383 0.09040974  1.8889983 110 0.06151704
3           cnsex -0.09466906 0.19431349 -0.4871976  89 0.62731001
4 Europe_vs_Others -0.05671159 0.22223781 -0.2551843  90 0.79916210
5        z_h3isei -0.15166447 0.10210395 -1.4853929  78 0.14146661
```

```
6          z_cngew -0.08057250 0.10178347 -0.7916069  79 0.43095108
7        z_son5mon  0.06308821 0.09863203  0.6396321  92 0.52399984
>
> # z_son5mot
> model3_mot <- with(imp, lm(z_son5mot ~ z_logMeth + cnsex + Europe_vs_Others +
 z_h3isei + z_cngew + z_son5mon))
> pooled_model3_mot <- pool(model3_mot)
> cat("\n========== MODEL 3: z_logMeth -> z_son5mot ==========\n")

========== MODEL 3: z_logMeth -> z_son5mot ==========
> summary_df3_mot <- summary(pooled_model3_mot, conf.int = FALSE)
> summary_df3_mot$df <- round(summary_df3_mot$df, 0)
> print(summary_df3_mot)
              term    estimate  std.error  statistic  df   p.value
1      (Intercept)  0.23665569 0.20374755  1.1615143 104 0.2480912
2        z_logMeth  0.07288522 0.08858746  0.8227487 114 0.4123760
3            cnsex -0.10737169 0.19396117 -0.5535731  86 0.5813149
4 Europe_vs_Others -0.26327240 0.22531981 -1.1684388  81 0.2460776
5         z_h3isei -0.16434829 0.10425493 -1.5764079  68 0.1195973
6          z_cngew -0.03506680 0.09457116 -0.3707981 103 0.7115483
7        z_son5mon -0.02563186 0.10290267 -0.2490884  73 0.8039952
>
> # z_son5koo
> model3_koo <- with(imp, lm(z_son5koo ~ z_logMeth + cnsex + Europe_vs_Others +
z_h3isei + z_cngew + z_son5mon))
> pooled_model3_koo <- pool(model3_koo)
> cat("\n========== MODEL 3: z_logMeth -> z_son5koo ==========\n")

========== MODEL 3: z_logMeth -> z_son5koo ==========
> summary_df3_koo <- summary(pooled_model3_koo, conf.int = FALSE)
> summary_df3_koo$df <- round(summary_df3_koo$df, 0)
> print(summary_df3_koo)
              term      estimate  std.error     statistic  df    p.value
1      (Intercept)  2.852497e-01 0.20273968  1.4069751705 108 0.16231392
2        z_logMeth  2.602287e-02 0.08863144  0.2936076404 115 0.76958751
3            cnsex -1.338104e-01 0.19217740 -0.6962859971  91 0.48802970
4 Europe_vs_Others -2.954740e-01 0.22722093 -1.3003817755  79 0.19724728
5         z_h3isei -1.678659e-01 0.09483928 -1.7700035649 104 0.07967101
6          z_cngew  5.582968e-05 0.09333581  0.0005981593 110 0.99952382
7        z_son5mon  6.394720e-02 0.10249788  0.6238880742  75 0.53458691
>
> # ============================================================
> # MODEL 4: cngrup -> z_logMeth
> # Covariates: cnsex, Europe_vs_Others, z_h3isei, z_cngew, z_h3tage, z_cnrandalt,
cnerr
> # ============================================================
>
> model4 <- with(imp, lm(z_logMeth ~ cngrup + cnsex + Europe_vs_Others + z_h3isei +
z_cngew +
+                            z_h3tage + z_cnrandalt + cnerr))
> pooled_model4 <- pool(model4)
> cat("\n========== MODEL 4 ==========\n")
```

```
========== MODEL 4 ==========
> summary_df4 <- summary(pooled_model4, conf.int = FALSE)
> summary_df4$df <- round(summary_df4$df, 0)
> print(summary_df4)
              term    estimate  std.error  statistic  df     p.value
1      (Intercept) -0.34762859 0.30865789 -1.1262586 121 0.262287301
2           cngrup -0.37473970 0.18118304 -2.0682935 121 0.040743831
3            cnsex  0.15524601 0.17318268  0.8964292 121 0.371803560
4 Europe_vs_Others  0.07975133 0.20309818  0.3926738 121 0.695252850
5         z_h3isei -0.05535785 0.09315197 -0.5942747 116 0.553486272
6          z_cngew -0.04508678 0.08871194 -0.5082380 121 0.612213925
7         z_h3tage -0.13816032 0.08983447 -1.5379433 121 0.126677594
8      z_cnrandalt  0.23679854 0.08968717  2.6402723 121 0.009376625
9            cnerr  0.46509145 0.26989974  1.7232008 121 0.087406706
>
>
> # ============================================================
> # MODELS 5-7: Simple Mediations
> # ============================================================
>
> library(mice)
> library(mediation)
>
> # Load transformed dataset
> imp <- readRDS("imputation_20_PMM_h3isei_transformed.rds")
>
> # ============================================================
> # MODEL 5: cngrup -> z_logMeth -> z_son5kon
> # ============================================================
>
> run_mediation5 <- function(data) {
+   med_model <- lm(z_logMeth ~ cngrup + cnerr + z_cnrandalt + z_h3tage +
+                     cnsex + Europe_vs_Others + z_cngew + z_h3isei,
+                   data = data)
+   out_model <- lm(z_son5kon ~ cngrup + z_logMeth + cnerr + z_cnrandalt +
+                     cnsex + z_son5mon + Europe_vs_Others + z_h3isei + z_cngew,
+                   data = data)
+   result <- mediate(med_model, out_model, treat = "cngrup", mediator = "z_logMeth",
+                     sims = 500, boot = TRUE)
+   data.frame(acme = result$d0, ade = result$z0, total = result$tau.coef,
+              prop = result$n0, p_acme = result$d0.p, p_ade = result$z0.p)
+ }
>
> set.seed(2026)
> results_list5 <- list()
> for(i in 1:imp$m) {
+   cat("Processing imputation", i, "of", imp$m, " (Model 5)\n")
+   data_i <- complete(imp, action = i)
+   results_list5[[i]] <- run_mediation5(data_i)
+ }
Processing imputation 1 of 20  (Model 5)
Running nonparametric bootstrapProcessing imputation 2 of 20  (Model 5)
Running nonparametric bootstrapProcessing imputation 3 of 20  (Model 5)
```

```
Running nonparametric bootstrapProcessing imputation 4 of 20  (Model 5)
Running nonparametric bootstrapProcessing imputation 5 of 20  (Model 5)
Running nonparametric bootstrapProcessing imputation 6 of 20  (Model 5)
Running nonparametric bootstrapProcessing imputation 7 of 20  (Model 5)
Running nonparametric bootstrapProcessing imputation 8 of 20  (Model 5)
Running nonparametric bootstrapProcessing imputation 9 of 20  (Model 5)
Running nonparametric bootstrapProcessing imputation 10 of 20  (Model 5)
Running nonparametric bootstrapProcessing imputation 11 of 20  (Model 5)
Running nonparametric bootstrapProcessing imputation 12 of 20  (Model 5)
Running nonparametric bootstrapProcessing imputation 13 of 20  (Model 5)
Running nonparametric bootstrapProcessing imputation 14 of 20  (Model 5)
Running nonparametric bootstrapProcessing imputation 15 of 20  (Model 5)
Running nonparametric bootstrapProcessing imputation 16 of 20  (Model 5)
Running nonparametric bootstrapProcessing imputation 17 of 20  (Model 5)
Running nonparametric bootstrapProcessing imputation 18 of 20  (Model 5)
Running nonparametric bootstrapProcessing imputation 19 of 20  (Model 5)
Running nonparametric bootstrapProcessing imputation 20 of 20  (Model 5)
Running nonparametric bootstrap> all_results5 <- do.call(rbind, results_list5)
>
> cat("\n========== MODEL 5: cngrup -> z_logMeth -> z_son5kon ==========\n")

========== MODEL 5: cngrup -> z_logMeth -> z_son5kon ==========
> cat("Indirect Effect (ACME):", round(mean(all_results5$acme), 3),
+     "p =", round(mean(all_results5$p_acme), 4), "\n")
Indirect Effect (ACME): -0.085 p = 0.0866
> cat("Direct Effect (ADE):", round(mean(all_results5$ade), 3),
+     "p =", round(mean(all_results5$p_ade), 4), "\n")
Direct Effect (ADE): -0.025 p = 0.7582
> cat("Total Effect:", round(mean(all_results5$total), 3), "\n")
Total Effect: -0.11
> cat("Proportion Mediated:", round(mean(all_results5$prop), 3), "\n")
Proportion Mediated: 1.47
>
> # ============================================================
> # MODEL 6: z_logMeth -> z_son5kon -> z_son5iq
> # ============================================================
>
> run_mediation6 <- function(data) {
+   med_model <- lm(z_son5kon ~ z_logMeth + z_h3tage + cnsex +
+                     Europe_vs_Others + z_cngew + z_h3isei + z_son5mon,
+                   data = data)
+   out_model <- lm(z_son5iq ~ z_logMeth + z_son5kon + cnsex +
+                     Europe_vs_Others + z_h3isei + z_cngew,
+                   data = data)
+   result <- mediate(med_model, out_model, treat = "z_logMeth", mediator = "z_son5kon",
+                     sims = 500, boot = TRUE)
+   data.frame(acme = result$d0, ade = result$z0, total = result$tau.coef,
+              prop = result$n0, p_acme = result$d0.p, p_ade = result$z0.p)
+ }
>
> set.seed(2026)
> results_list6 <- list()
> for(i in 1:imp$m) {
```

```
+   cat("Processing imputation", i, "of", imp$m, " (Model 6)\n")
+   data_i <- complete(imp, action = i)
+   results_list6[[i]] <- run_mediation6(data_i)
+ }
Processing imputation 1 of 20  (Model 6)
Running nonparametric bootstrapProcessing imputation 2 of 20  (Model 6)
Running nonparametric bootstrapProcessing imputation 3 of 20  (Model 6)
Running nonparametric bootstrapProcessing imputation 4 of 20  (Model 6)
Running nonparametric bootstrapProcessing imputation 5 of 20  (Model 6)
Running nonparametric bootstrapProcessing imputation 6 of 20  (Model 6)
Running nonparametric bootstrapProcessing imputation 7 of 20  (Model 6)
Running nonparametric bootstrapProcessing imputation 8 of 20  (Model 6)
Running nonparametric bootstrapProcessing imputation 9 of 20  (Model 6)
Running nonparametric bootstrapProcessing imputation 10 of 20  (Model 6)
Running nonparametric bootstrapProcessing imputation 11 of 20  (Model 6)
Running nonparametric bootstrapProcessing imputation 12 of 20  (Model 6)
Running nonparametric bootstrapProcessing imputation 13 of 20  (Model 6)
Running nonparametric bootstrapProcessing imputation 14 of 20  (Model 6)
Running nonparametric bootstrapProcessing imputation 15 of 20  (Model 6)
Running nonparametric bootstrapProcessing imputation 16 of 20  (Model 6)
Running nonparametric bootstrapProcessing imputation 17 of 20  (Model 6)
Running nonparametric bootstrapProcessing imputation 18 of 20  (Model 6)
Running nonparametric bootstrapProcessing imputation 19 of 20  (Model 6)
Running nonparametric bootstrapProcessing imputation 20 of 20  (Model 6)
Running nonparametric bootstrap> all_results6 <- do.call(rbind, results_list6)
>
> cat("\n========== MODEL 6: z_logMeth -> z_son5kon -> z_son5iq ==========\n")

========== MODEL 6: z_logMeth -> z_son5kon -> z_son5iq ==========
> cat("Indirect Effect (ACME):", round(mean(all_results6$acme), 3),
+     "p =", round(mean(all_results6$p_acme), 4), "\n")
Indirect Effect (ACME): -0.083 p = 0.0124
> cat("Direct Effect (ADE):", round(mean(all_results6$ade), 3),
+     "p =", round(mean(all_results6$p_ade), 4), "\n")
Direct Effect (ADE): 0.004 p = 0.8024
> cat("Total Effect:", round(mean(all_results6$total), 3), "\n")
Total Effect: -0.079
> cat("Proportion Mediated:", round(mean(all_results6$prop), 3), "\n")
Proportion Mediated: 1.151
>
> # ============================================================
> # MODEL 7: z_e3part_rev -> z_logMeth -> z_son5kon
> # (Create z_e3part_rev directly from transformed data)
> # ============================================================
>
> library(mice)
> library(mediation)
>
> # Load transformed dataset (with z-scores)
> imp <- readRDS("imputation_20_PMM_h3isei_transformed.rds")
>
> # ============================================================
> # Create z_e3part_rev on the fly
```

```
> # ============================================================
>
> # Extract long format and create reversed variable
> imp_long <- complete(imp, action = "long", include = TRUE)
>
> # Reverse e3part: higher values = more disagreement
> imp_long$z_e3part_rev <- -imp_long$z_e3part
>
> # Convert back to mids object
> imp_rev <- as.mids(imp_long)
>
> # Optional: save it for later use
> saveRDS(imp_rev, file = "imputation_20_PMM_h3isei_reversed.rds")
>
> # ============================================================
> # Run Mediation Model 7
> # ============================================================
>
> run_mediation7 <- function(data) {
+
+   med_model <- lm(z_logMeth ~ z_e3part_rev + z_h3tage + cnsex +
+                     Europe_vs_Others + z_cngew + z_h3isei,
+                   data = data)
+
+   out_model <- lm(z_son5kon ~ z_e3part_rev + z_logMeth + cnsex +
+                     z_son5mon + Europe_vs_Others + z_h3isei + z_cngew,
+                   data = data)
+
+   result <- mediate(med_model, out_model,
+                     treat = "z_e3part_rev",
+                     mediator = "z_logMeth",
+                     sims = 500,
+                     boot = TRUE)
+
+   data.frame(
+     acme = result$d0,
+     ade = result$z0,
+     total = result$tau.coef,
+     prop = result$n0,
+     p_acme = result$d0.p,
+     p_ade = result$z0.p
+   )
+ }
>
> # Run on all imputations
> set.seed(2026)
> results_list7 <- list()
>
> for(i in 1:imp_rev$m) {
+   cat("Processing imputation", i, "of", imp_rev$m, " (Model 7)\n")
+   data_i <- complete(imp_rev, action = i)
+   results_list7[[i]] <- run_mediation7(data_i)
+ }
```

```
Processing imputation 1 of 20  (Model 7)
Running nonparametric bootstrapProcessing imputation 2 of 20  (Model 7)
Running nonparametric bootstrapProcessing imputation 3 of 20  (Model 7)
Running nonparametric bootstrapProcessing imputation 4 of 20  (Model 7)
Running nonparametric bootstrapProcessing imputation 5 of 20  (Model 7)
Running nonparametric bootstrapProcessing imputation 6 of 20  (Model 7)
Running nonparametric bootstrapProcessing imputation 7 of 20  (Model 7)
Running nonparametric bootstrapProcessing imputation 8 of 20  (Model 7)
Running nonparametric bootstrapProcessing imputation 9 of 20  (Model 7)
Running nonparametric bootstrapProcessing imputation 10 of 20  (Model 7)
Running nonparametric bootstrapProcessing imputation 11 of 20  (Model 7)
Running nonparametric bootstrapProcessing imputation 12 of 20  (Model 7)
Running nonparametric bootstrapProcessing imputation 13 of 20  (Model 7)
Running nonparametric bootstrapProcessing imputation 14 of 20  (Model 7)
Running nonparametric bootstrapProcessing imputation 15 of 20  (Model 7)
Running nonparametric bootstrapProcessing imputation 16 of 20  (Model 7)
Running nonparametric bootstrapProcessing imputation 17 of 20  (Model 7)
Running nonparametric bootstrapProcessing imputation 18 of 20  (Model 7)
Running nonparametric bootstrapProcessing imputation 19 of 20  (Model 7)
Running nonparametric bootstrapProcessing imputation 20 of 20  (Model 7)
Running nonparametric bootstrap>
> # Combine and pool results
> all_results7 <- do.call(rbind, results_list7)
>
> cat("\n========== MODEL 7: z_e3part_rev -> z_logMeth -> z_son5kon ==========\n")

========== MODEL 7: z_e3part_rev -> z_logMeth -> z_son5kon ==========
> cat("Indirect Effect (ACME):", round(mean(all_results7$acme), 3),
+     " (SD =", round(sd(all_results7$acme), 3), ")\n")
Indirect Effect (ACME): 0.064  (SD = 0.007 )
> cat("p-value:", round(mean(all_results7$p_acme), 4), "\n\n")
p-value: 0.023

> cat("Direct Effect (ADE):", round(mean(all_results7$ade), 3),
+     " (SD =", round(sd(all_results7$ade), 3), ")\n")
Direct Effect (ADE): 0.051  (SD = 0.05 )
> cat("p-value:", round(mean(all_results7$p_ade), 4), "\n\n")
p-value: 0.6024

> cat("Total Effect:", round(mean(all_results7$total), 3), "\n")
Total Effect: 0.115
> cat("Proportion Mediated:", round(mean(all_results7$prop), 3), "\n")
Proportion Mediated: 0.677
>
>
> # ============================================================
> # MODEL 8: Moderated Mediation (IG vs CG)
> # z_e3part_rev -> z_logMeth -> z_son5kon
> # ============================================================
>
> library(mice)
> library(mediation)
>
```

```
> # Load transformed dataset and create imp_rev (if not already in memory)
> imp <- readRDS("imputation_20_PMM_h3isei_transformed.rds")
>
> imp_long <- complete(imp, action = "long", include = TRUE)
> imp_long$z_e3part_rev <- -imp_long$z_e3part
> imp_rev <- as.mids(imp_long)
>
> # ============================================================
> # Function for one group
> # ============================================================
>
> run_mediation_group <- function(data, group_value) {
+
+   data_group <- data[data$cngrup == group_value, ]
+
+   if(nrow(data_group) < 20) {
+     return(data.frame(acme = NA, p_acme = NA, n = nrow(data_group)))
+   }
+
+   med_model <- lm(z_logMeth ~ z_e3part_rev + z_h3tage + cnsex +
+                     Europe_vs_Others + z_cngew + z_h3isei + cnerr + z_cnrandalt,
+                   data = data_group)
+
+   out_model <- lm(z_son5kon ~ z_e3part_rev + z_logMeth + cnsex +
+                     z_son5mon + Europe_vs_Others + z_h3isei + z_cngew,
+                   data = data_group)
+
+   result <- mediate(med_model, out_model,
+                     treat = "z_e3part_rev",
+                     mediator = "z_logMeth",
+                     sims = 200,
+                     boot = TRUE)
+
+   return(data.frame(acme = result$d0, p_acme = result$d0.p, n = nrow(data_group)))
+ }
>
> # ============================================================
> # Run on all imputations
> # ============================================================
>
> set.seed(2026)
>
> results_cg <- list()
> results_ig <- list()
>
> for(i in 1:imp_rev$m) {
+   cat("Imputation", i, "of", imp_rev$m, "\n")
+   data_i <- complete(imp_rev, action = i)
+
+   results_cg[[i]] <- run_mediation_group(data_i, group_value = 0)
+   results_ig[[i]] <- run_mediation_group(data_i, group_value = 1)
+ }
Imputation 1 of 20
```

```
Running nonparametric bootstrap

 Running nonparametric bootstrapImputation 2 of 20
Running nonparametric bootstrap

 Running nonparametric bootstrapImputation 3 of 20
Running nonparametric bootstrap

 Running nonparametric bootstrapImputation 4 of 20
Running nonparametric bootstrap

 Running nonparametric bootstrapImputation 5 of 20
Running nonparametric bootstrap

 Running nonparametric bootstrapImputation 6 of 20
Running nonparametric bootstrap

 Running nonparametric bootstrapImputation 7 of 20
Running nonparametric bootstrap

 Running nonparametric bootstrapImputation 8 of 20
Running nonparametric bootstrap

 Running nonparametric bootstrapImputation 9 of 20
Running nonparametric bootstrap

 Running nonparametric bootstrapImputation 10 of 20
Running nonparametric bootstrap

 Running nonparametric bootstrapImputation 11 of 20
Running nonparametric bootstrap

 Running nonparametric bootstrapImputation 12 of 20
Running nonparametric bootstrap

 Running nonparametric bootstrapImputation 13 of 20
Running nonparametric bootstrap
```

```
 Running nonparametric bootstrapImputation 14 of 20
Running nonparametric bootstrap

 Running nonparametric bootstrapImputation 15 of 20
Running nonparametric bootstrap

 Running nonparametric bootstrapImputation 16 of 20
Running nonparametric bootstrap

 Running nonparametric bootstrapImputation 17 of 20
Running nonparametric bootstrap

 Running nonparametric bootstrapImputation 18 of 20
Running nonparametric bootstrap

 Running nonparametric bootstrapImputation 19 of 20
Running nonparametric bootstrap

 Running nonparametric bootstrapImputation 20 of 20
Running nonparametric bootstrap

Running nonparametric bootstrap

 There were 50 or more warnings (use warnings() to see the first 50)>
 > # ============================================================
 > # Combine and pool results
 > # ============================================================
 >
 > cg_all <- do.call(rbind, results_cg)
 > ig_all <- do.call(rbind, results_ig)
 >
 > # Remove NA
 > cg_clean <- cg_all[!is.na(cg_all$acme), ]
 > ig_clean <- ig_all[!is.na(ig_all$acme), ]
 >
 > # Control Group
 > pooled_acme_cg <- mean(cg_clean$acme)
 > pooled_p_cg <- mean(cg_clean$p_acme)
 > sd_acme_cg <- sd(cg_clean$acme)
 > n_cg <- round(mean(cg_clean$n))
 >
 > # Intervention Group
 > pooled_acme_ig <- mean(ig_clean$acme)
 > pooled_p_ig <- mean(ig_clean$p_acme)
```

```
> sd_acme_ig <- sd(ig_clean$acme)
> n_ig <- round(mean(ig_clean$n))
>
> # Difference
> diff_acme <- pooled_acme_ig - pooled_acme_cg
> se_diff <- sqrt(sd_acme_cg^2 + sd_acme_ig^2)
> z_diff <- diff_acme / se_diff
> p_diff <- 2 * (1 - pnorm(abs(z_diff)))
>
> # ============================================================
> # Print results
> # ============================================================
>
> cat("\n============================================================\n")

============================================================
> cat("MODEL 8: Moderated Mediation\n")
MODEL 8: Moderated Mediation
> cat("z_e3part_rev -> z_logMeth -> z_son5kon\n")
z_e3part_rev -> z_logMeth -> z_son5kon
> cat("============================================================\n\n")
============================================================

>
> cat("Control Group (CG):\n")
Control Group (CG):
> cat("  N:", n_cg, "\n")
  N: 60
> cat("  Indirect effect:", round(pooled_acme_cg, 4), "\n")
  Indirect effect: 0.0803
> cat("  SD:", round(sd_acme_cg, 4), "\n")
  SD: 0.0252
> cat("  p-value:", round(pooled_p_cg, 4), "\n\n")
  p-value: 0.265

>
> cat("Intervention Group (IG):\n")
Intervention Group (IG):
> cat("  N:", n_ig, "\n")
  N: 72
> cat("  Indirect effect:", round(pooled_acme_ig, 4), "\n")
  Indirect effect: 0.0639
> cat("  SD:", round(sd_acme_ig, 4), "\n")
  SD: 0.0068
> cat("  p-value:", round(pooled_p_ig, 4), "\n\n")
  p-value: 0.0875

>
> cat("Difference (IG - CG):", round(diff_acme, 4), "\n")
Difference (IG - CG): -0.0164
> cat("  SE:", round(se_diff, 4), "\n")
  SE: 0.0261
> cat("  z-value:", round(z_diff, 2), "\n")
```

```
  z-value: -0.63
> cat("  p-value:", round(p_diff, 4), "\n\n")
  p-value: 0.531

>
> if(p_diff < 0.05) {
+   cat("✓ Significant moderation\n")
+ } else {
+   cat("✗ No significant moderation\n")
+ }
✗ No significant moderation
>
> # ============================================================
> # Save results
> # ============================================================
>
> group_results <- data.frame(
+   Group = c("Control (CG)", "Intervention (IG)"),
+   N = c(n_cg, n_ig),
+   Indirect_Effect = c(pooled_acme_cg, pooled_acme_ig),
+   SD = c(sd_acme_cg, sd_acme_ig),
+   p_value = c(pooled_p_cg, pooled_p_ig)
+ )
>
> write.csv(group_results, "model8_moderated_mediation_h3isei.csv", row.names = FALSE)
>
> cat("\nResults saved to model8_moderated_mediation_h3isei.csv\n")

Results saved to model8_moderated_mediation_h3isei.csv
>
>
> # ============================================================
> # MODEL 9: Serial Mediation
> # z_e3part_rev -> z_logMeth -> z_son5kon -> z_son5iq
> # ============================================================
>
> library(mice)
> library(boot)
>
> # Load transformed dataset and create imp_rev (if not already in memory)
> imp <- readRDS("imputation_20_PMM_h3isei_transformed.rds")
>
> imp_long <- complete(imp, action = "long", include = TRUE)
> imp_long$z_e3part_rev <- -imp_long$z_e3part
> imp_rev <- as.mids(imp_long)
>
> # ============================================================
> # Function to run serial mediation on one dataset
> # ============================================================
>
> run_serial_mediation <- function(data) {
+
+   # Path a: z_e3part_rev -> z_logMeth
```

```
+   model_a <- lm(z_logMeth ~ z_e3part_rev + cnsex + z_h3tage +
+                   Europe_vs_Others + z_cngew + z_h3isei,
+                 data = data)
+   a_coef <- coef(model_a)["z_e3part_rev"]
+
+   # Path b1: z_logMeth -> z_son5kon
+   model_b1 <- lm(z_son5kon ~ z_logMeth + cnsex + z_son5mon +
+                    Europe_vs_Others + z_cngew + z_h3isei,
+                  data = data)
+   b1_coef <- coef(model_b1)["z_logMeth"]
+
+   # Path b2: z_son5kon -> z_son5iq
+   model_b2 <- lm(z_son5iq ~ z_son5kon + cnsex + Europe_vs_Others +
+                    z_cngew + z_h3isei,
+                  data = data)
+   b2_coef <- coef(model_b2)["z_son5kon"]
+
+   # Serial indirect effect
+   ab_serial <- a_coef * b1_coef * b2_coef
+
+   # Bootstrap for standard error
+   boot_serial <- function(d, indices) {
+     d_sub <- d[indices, ]
+     a_boot <- coef(lm(z_logMeth ~ z_e3part_rev + cnsex + z_h3tage +
+                         Europe_vs_Others + z_cngew + z_h3isei, data = d_sub))
["z_e3part_rev"]
+     b1_boot <- coef(lm(z_son5kon ~ z_logMeth + cnsex + z_son5mon +
+                          Europe_vs_Others + z_cngew + z_h3isei, data = d_sub))
["z_logMeth"]
+     b2_boot <- coef(lm(z_son5iq ~ z_son5kon + cnsex + Europe_vs_Others +
+                          z_cngew + z_h3isei, data = d_sub))["z_son5kon"]
+     return(a_boot * b1_boot * b2_boot)
+   }
+
+   boot_result <- boot::boot(data, boot_serial, R = 200)
+   boot_se <- sd(boot_result$t, na.rm = TRUE)
+   p_serial <- 2 * (1 - pnorm(abs(ab_serial / boot_se)))
+
+   return(data.frame(
+     a = a_coef,
+     b1 = b1_coef,
+     b2 = b2_coef,
+     ab_serial = ab_serial,
+     boot_se = boot_se,
+     p = p_serial
+   ))
+ }
>
> # ==============================================================
> # Run on all imputations
> # ==============================================================
>
> set.seed(2026)
```

```
> serial_results <- list()
>
> for(i in 1:imp_rev$m) {
+   cat("Processing imputation", i, "of", imp_rev$m, " (Model 9)\n")
+   data_i <- complete(imp_rev, action = i)
+   serial_results[[i]] <- run_serial_mediation(data_i)
+ }
Processing imputation 1 of 20  (Model 9)
Processing imputation 2 of 20  (Model 9)
Processing imputation 3 of 20  (Model 9)
Processing imputation 4 of 20  (Model 9)
Processing imputation 5 of 20  (Model 9)
Processing imputation 6 of 20  (Model 9)
Processing imputation 7 of 20  (Model 9)
Processing imputation 8 of 20  (Model 9)
Processing imputation 9 of 20  (Model 9)
Processing imputation 10 of 20  (Model 9)
Processing imputation 11 of 20  (Model 9)
Processing imputation 12 of 20  (Model 9)
Processing imputation 13 of 20  (Model 9)
Processing imputation 14 of 20  (Model 9)
Processing imputation 15 of 20  (Model 9)
Processing imputation 16 of 20  (Model 9)
Processing imputation 17 of 20  (Model 9)
Processing imputation 18 of 20  (Model 9)
Processing imputation 19 of 20  (Model 9)
Processing imputation 20 of 20  (Model 9)
>
> # Combine results
> all_serial <- do.call(rbind, serial_results)
>
> # ============================================================
> # Print results
> # ============================================================
>
> cat("\n============================================================\n")

============================================================
> cat("MODEL 9: Serial Mediation\n")
MODEL 9: Serial Mediation
> cat("z_e3part_rev -> z_logMeth -> z_son5kon -> z_son5iq\n")
z_e3part_rev -> z_logMeth -> z_son5kon -> z_son5iq
> cat("============================================================\n\n")
============================================================

>
> cat("Path a (z_e3part_rev -> z_logMeth):", round(mean(all_serial$a), 4), "\n")
Path a (z_e3part_rev -> z_logMeth): 0.276
> cat("Path b1 (z_logMeth -> z_son5kon):", round(mean(all_serial$b1), 4), "\n")
Path b1 (z_logMeth -> z_son5kon): 0.2441
> cat("Path b2 (z_son5kon -> z_son5iq):", round(mean(all_serial$b2), 4), "\n\n")
Path b2 (z_son5kon -> z_son5iq): -0.3583
```

```
>
> cat("Serial Indirect Effect (a * b1 * b2):\n")
Serial Indirect Effect (a * b1 * b2):
> cat("  Estimate:", round(mean(all_serial$ab_serial), 4), "\n")
  Estimate: -0.0241
> cat("  Boot SE:", round(mean(all_serial$boot_se), 4), "\n")
  Boot SE: 0.0152
> cat("  p-value:", round(mean(all_serial$p), 4), "\n\n")
  p-value: 0.119

>
> if(mean(all_serial$p) < 0.05) {
+   cat("✓ The serial mediation effect is significant.\n")
+ } else {
+   cat("✗ The serial mediation effect is not significant.\n")
+ }
✗ The serial mediation effect is not significant.
>
> # ============================================================
> # Save results
> # ============================================================
>
> write.csv(all_serial, "model9_serial_mediation_h3isei.csv", row.names = FALSE)
>
> cat("\nResults saved to model9_serial_mediation_h3isei.csv\n")

Results saved to model9_serial_mediation_h3isei.csv
>
>
> # ============================================================
> # MODEL 10: Moderated Serial Mediation (IG vs CG)
> # z_e3part_rev -> z_logMeth -> z_son5kon -> z_son5iq
> # ============================================================
>
> library(mice)
> library(boot)
>
> # Load transformed dataset and create imp_rev (if not already in memory)
> imp <- readRDS("imputation_20_PMM_h3isei_transformed.rds")
>
> imp_long <- complete(imp, action = "long", include = TRUE)
> imp_long$z_e3part_rev <- -imp_long$z_e3part
> imp_rev <- as.mids(imp_long)
>
> # ============================================================
> # Function for one group
> # ============================================================
>
> run_serial_group <- function(data, group_value, group_name) {
+
+   data_group <- data[data$cngrup == group_value, ]
+
+   cat("  ", group_name, "N =", nrow(data_group), "\n")
```

```
+
+   if(nrow(data_group) < 20) {
+     return(data.frame(ab_serial = NA, p = NA, n = nrow(data_group)))
+   }
+
+   # Path a
+   model_a <- lm(z_logMeth ~ z_e3part_rev + cnsex + z_h3tage +
+                   Europe_vs_Others + z_cngew + z_h3isei,
+                 data = data_group)
+   a_coef <- coef(model_a)["z_e3part_rev"]
+
+   # Path b1
+   model_b1 <- lm(z_son5kon ~ z_logMeth + cnsex + z_son5mon +
+                    Europe_vs_Others + z_cngew + z_h3isei,
+                  data = data_group)
+   b1_coef <- coef(model_b1)["z_logMeth"]
+
+   # Path b2
+   model_b2 <- lm(z_son5iq ~ z_son5kon + cnsex + Europe_vs_Others +
+                    z_cngew + z_h3isei,
+                  data = data_group)
+   b2_coef <- coef(model_b2)["z_son5kon"]
+
+   # Serial indirect
+   ab_serial <- a_coef * b1_coef * b2_coef
+
+   # Bootstrap
+   boot_serial <- function(d, indices) {
+     d_sub <- d[indices, ]
+     a_boot <- coef(lm(z_logMeth ~ z_e3part_rev + cnsex + z_h3tage +
+                         Europe_vs_Others + z_cngew + z_h3isei, data = d_sub))
["z_e3part_rev"]
+     b1_boot <- coef(lm(z_son5kon ~ z_logMeth + cnsex + z_son5mon +
+                          Europe_vs_Others + z_cngew + z_h3isei, data = d_sub))
["z_logMeth"]
+     b2_boot <- coef(lm(z_son5iq ~ z_son5kon + cnsex + Europe_vs_Others +
+                          z_cngew + z_h3isei, data = d_sub))["z_son5kon"]
+     return(a_boot * b1_boot * b2_boot)
+   }
+
+   boot_result <- tryCatch({
+     boot::boot(data_group, boot_serial, R = 200)
+   }, error = function(e) return(NULL))
+
+   if(!is.null(boot_result)) {
+     boot_se <- sd(boot_result$t, na.rm = TRUE)
+     p_serial <- 2 * (1 - pnorm(abs(ab_serial / boot_se)))
+   } else {
+     p_serial <- NA
+   }
+
+   return(data.frame(
+     group = group_name,
```

```
+     n = nrow(data_group),
+     ab_serial = ab_serial,
+     p = p_serial
+   ))
+ }
>
> # ============================================================
> # Run on all imputations
> # ============================================================
>
> set.seed(2026)
>
> results_cg <- list()
> results_ig <- list()
>
> for(i in 1:imp_rev$m) {
+   cat("\nImputation", i, "of", imp_rev$m, "\n")
+   data_i <- complete(imp_rev, action = i)
+
+   results_cg[[i]] <- run_serial_group(data_i, group_value = 0, group_name = "CG")
+   results_ig[[i]] <- run_serial_group(data_i, group_value = 1, group_name = "IG")
+ }

Imputation 1 of 20
   CG N = 60
   IG N = 72

Imputation 2 of 20
   CG N = 60
   IG N = 72

Imputation 3 of 20
   CG N = 60
   IG N = 72

Imputation 4 of 20
   CG N = 60
   IG N = 72

Imputation 5 of 20
   CG N = 60
   IG N = 72

Imputation 6 of 20
   CG N = 60
   IG N = 72

Imputation 7 of 20
   CG N = 60
   IG N = 72

Imputation 8 of 20
   CG N = 60
```

```
   IG N = 72

Imputation 9 of 20
   CG N = 60
   IG N = 72

Imputation 10 of 20
   CG N = 60
   IG N = 72

Imputation 11 of 20
   CG N = 60
   IG N = 72

Imputation 12 of 20
   CG N = 60
   IG N = 72

Imputation 13 of 20
   CG N = 60
   IG N = 72

Imputation 14 of 20
   CG N = 60
   IG N = 72

Imputation 15 of 20
   CG N = 60
   IG N = 72

Imputation 16 of 20
   CG N = 60
   IG N = 72

Imputation 17 of 20
   CG N = 60
   IG N = 72

Imputation 18 of 20
   CG N = 60
   IG N = 72

Imputation 19 of 20
   CG N = 60
   IG N = 72

Imputation 20 of 20
   CG N = 60
   IG N = 72
>
> # ============================================================
> # Combine results
> # ============================================================
```

```
>
> cg_all <- do.call(rbind, results_cg)
> ig_all <- do.call(rbind, results_ig)
>
> # Remove NA
> cg_clean <- cg_all[!is.na(cg_all$ab_serial), ]
> ig_clean <- ig_all[!is.na(ig_all$ab_serial), ]
>
> # Pool within groups
> ab_cg <- mean(cg_clean$ab_serial)
> p_cg <- mean(cg_clean$p)
> n_cg <- round(mean(cg_clean$n))
> sd_cg <- sd(cg_clean$ab_serial)
>
> ab_ig <- mean(ig_clean$ab_serial)
> p_ig <- mean(ig_clean$p)
> n_ig <- round(mean(ig_clean$n))
> sd_ig <- sd(ig_clean$ab_serial)
>
> # Difference between groups
> diff_serial <- ab_ig - ab_cg
> se_diff <- sqrt(sd_cg^2 + sd_ig^2)
> z_diff <- diff_serial / se_diff
> p_diff <- 2 * (1 - pnorm(abs(z_diff)))
>
> # ============================================================
> # Print results
> # ============================================================
>
> cat("\n============================================================\n")

============================================================
> cat("MODEL 10: Moderated Serial Mediation\n")
MODEL 10: Moderated Serial Mediation
> cat("z_e3part_rev -> z_logMeth -> z_son5kon -> z_son5iq\n")
z_e3part_rev -> z_logMeth -> z_son5kon -> z_son5iq
> cat("============================================================\n\n")
============================================================

>
> cat("Control Group (CG):\n")
Control Group (CG):
> cat("  N:", n_cg, "\n")
  N: 60
> cat("  Serial indirect effect:", round(ab_cg, 4), "\n")
  Serial indirect effect: -0.0298
> cat("  SD:", round(sd_cg, 4), "\n")
  SD: 0.0056
> cat("  p-value:", round(p_cg, 4), "\n\n")
  p-value: 0.347

>
> cat("Intervention Group (IG):\n")
```

```
Intervention Group (IG):
> cat("  N:", n_ig, "\n")
  N: 72
> cat("  Serial indirect effect:", round(ab_ig, 4), "\n")
  Serial indirect effect: -0.019
> cat("  SD:", round(sd_ig, 4), "\n")
  SD: 0.0032
> cat("  p-value:", round(p_ig, 4), "\n\n")
  p-value: 0.2145

>
> cat("Difference (IG - CG):", round(diff_serial, 4), "\n")
Difference (IG - CG): 0.0108
> cat("  SE:", round(se_diff, 4), "\n")
  SE: 0.0064
> cat("  z-value:", round(z_diff, 2), "\n")
  z-value: 1.67
> cat("  p-value:", round(p_diff, 4), "\n\n")
  p-value: 0.0953

>
> if(p_diff < 0.05) {
+   cat("✓ Significant moderation: The serial mediation effect differs between groups
.\n")
+ } else {
+   cat("✗ No significant moderation: The serial mediation effect does not differ
between groups.\n")
+ }
✗ No significant moderation: The serial mediation effect does not differ between
groups.
```

**Complete cases analyses**

```
# ============================================================
> # COMPLETE CASES ANALYSIS (NO IMPUTATION)
> # ============================================================
>
> library(dplyr)
> library(mediation)
> library(boot)
>
> # Load raw data
> raw_data <- read.csv("original dataset.csv")
>
> # ============================================================
> # Step 1: Convert dummy codes to NA
> # ============================================================
>
> dummy_codes <- c(-333, -444, -555, -777, -999)
>
> data_clean <- raw_data
>
> for(code in dummy_codes) {
+   data_clean[data_clean == code] <- NA
+ }
>
```

```
> # ============================================================
> # Step 2: Select variables and create complete cases dataset
> # ============================================================
>
> data_selected <- data_clean %>%
+   dplyr::select(cngrup, cnsex, h3tage, cnrandalt, h3isei, cngew,
+                 Europe_vs_Others, cnerr, logMeth, e3part,
+                 son5iq, son5kon, son5ver, son5mot, son5koo, son5mon)
>
> # Remove rows with any missing values
> data_complete <- na.omit(data_selected)
>
> cat("Original N:", nrow(data_selected), "\n")
Original N: 132
> cat("Complete cases N:", nrow(data_complete), "\n")
Complete cases N: 86
> cat("Rows removed:", nrow(data_selected) - nrow(data_complete), "\n")
Rows removed: 46
>
> # ============================================================
> # Step 3: Create z-scores for continuous variables
> # ============================================================
>
> data_complete <- data_complete %>%
+   mutate(
+     z_logMeth = as.numeric(scale(logMeth)),
+     z_h3isei = as.numeric(scale(h3isei)),  # <--- CHANGED: was z_hisei
+     z_cngew = as.numeric(scale(cngew)),
+     z_h3tage = as.numeric(scale(h3tage)),
+     z_cnrandalt = as.numeric(scale(cnrandalt)),
+     z_son5iq = as.numeric(scale(son5iq)),
+     z_son5kon = as.numeric(scale(son5kon)),
+     z_son5ver = as.numeric(scale(son5ver)),
+     z_son5mot = as.numeric(scale(son5mot)),
+     z_son5koo = as.numeric(scale(son5koo)),
+     z_son5mon = as.numeric(scale(son5mon)),
+     z_e3part = as.numeric(scale(e3part))
+   )
>
> # Create reversed e3part (higher = more disagreement)
> data_complete$z_e3part_rev <- -data_complete$z_e3part
>
> cat("\nZ-scores created with h3isei.\n")

Z-scores
> cat("Final N for all models:", nrow(data_complete), "\n")
Final N for all models: 86
>
> # ============================================================
> # MODEL 1: z_logMeth -> z_son5iq
> # Covariates: cnsex, Europe_vs_Others, z_h3isei, z_cngew
> # ============================================================
>
> model1 <- lm(z_son5iq ~ z_logMeth + cnsex + Europe_vs_Others + z_h3isei +
z_cngew,
+              data = data_complete)
>
> cat("\n========== MODEL 1 ==========\n")

========== MODEL 1 ==========
> summary(model1)

Call:
lm(formula = z_son5iq ~ z_logMeth + cnsex + Europe_vs_Others +
```

```
    z_h3isei + z_cngew, data = data_complete)

Residuals:
     Min       1Q   Median       3Q      Max 
-2.28810 -0.58020 -0.04377  0.62783  1.95005 

Coefficients:
                 Estimate Std. Error t value Pr(>|t|)  
(Intercept)       -0.1285     0.2238  -0.574   0.5673  
z_logMeth         -0.1100     0.1025  -1.073   0.2863  
cnsex              0.4939     0.2096   2.357   0.0209 *
Europe_vs_Others  -0.2274     0.2314  -0.983   0.3288  
z_h3isei           0.2556     0.1045   2.446   0.0166 *
z_cngew            0.2297     0.1067   2.153   0.0344 *
---
Signif. codes:  0 ‘***’ 0.001 ‘**’ 0.01 ‘*’ 0.05 ‘.’ 0.1 ‘ ’ 1

Residual standard error: 0.9373 on 80 degrees of freedom
Multiple R-squared:  0.1731,	Adjusted R-squared:  0.1214 
F-statistic: 3.348 on 5 and 80 DF,  p-value: 0.008496


> 
> # ============================================================
> # MODEL 2: z_logMeth -> z_son5kon
> # Covariates: cnsex, Europe_vs_Others, z_h3isei, z_cngew, z_son5mon
> # ============================================================
> 
> model2 <- lm(z_son5kon ~ z_logMeth + cnsex + Europe_vs_Others + z_h3isei + 
z_cngew + z_son5mon,
+              data = data_complete)
> 
> cat("\n========== MODEL 2 ==========\n")

========== MODEL 2 ==========
> summary(model2)

Call:
lm(formula = z_son5kon ~ z_logMeth + cnsex + Europe_vs_Others + 
    z_h3isei + z_cngew + z_son5mon, data = data_complete)

Residuals:
    Min      1Q  Median      3Q     Max 
-1.4441 -0.7486 -0.1339  0.6262  2.6390 

Coefficients:
                 Estimate Std. Error t value Pr(>|t|)   
(Intercept)       0.13461    0.22671   0.594  0.55438   
z_logMeth         0.31450    0.10258   3.066  0.00297 **
cnsex            -0.45636    0.20987  -2.174  0.03266 * 
Europe_vs_Others  0.18736    0.23598   0.794  0.42959   
z_h3isei         -0.08758    0.10521  -0.832  0.40764   
z_cngew           0.11968    0.10951   1.093  0.27776   
z_son5mon         0.11449    0.10561   1.084  0.28164   
---
Signif. codes:  0 ‘***’ 0.001 ‘**’ 0.01 ‘*’ 0.05 ‘.’ 0.1 ‘ ’ 1

Residual standard error: 0.9383 on 79 degrees of freedom
Multiple R-squared:  0.1818,	Adjusted R-squared:  0.1197 
F-statistic: 2.926 on 6 and 79 DF,  p-value: 0.01251


> 
> # ============================================================
> # MODEL 3: z_logMeth -> z_son5ver, z_son5mot, z_son5koo
> # Covariates: cnsex, Europe_vs_Others, z_h3isei, z_cngew, z_son5mon
```

```
> # ============================================================
>
> model3_ver <- lm(z_son5ver ~ z_logMeth + cnsex + Europe_vs_Others +
z_h3isei + z_cngew + z_son5mon,
+                  data = data_complete)
> model3_mot <- lm(z_son5mot ~ z_logMeth + cnsex + Europe_vs_Others +
z_h3isei + z_cngew + z_son5mon,
+                  data = data_complete)
> model3_koo <- lm(z_son5koo ~ z_logMeth + cnsex + Europe_vs_Others +
z_h3isei + z_cngew + z_son5mon,
+                  data = data_complete)
>
> cat("\n========== MODEL 3a: z_logMeth -> z_son5ver ==========\n")

========== MODEL 3a: z_logMeth -> z_son5ver ==========
> summary(model3_ver)

Call:
lm(formula = z_son5ver ~ z_logMeth + cnsex + Europe_vs_Others +
    z_h3isei + z_cngew + z_son5mon, data = data_complete)

Residuals:
    Min      1Q  Median      3Q     Max
-1.3291 -0.4483 -0.2758 -0.0616  3.4007

Coefficients:
                  Estimate Std. Error t value Pr(>|t|)
(Intercept)       0.006657   0.238678   0.028    0.978
z_logMeth         0.203627   0.107996   1.886    0.063 .
cnsex            -0.063299   0.220949  -0.286    0.775
Europe_vs_Others  0.043208   0.248431   0.174    0.862
z_h3isei         -0.159480   0.110761  -1.440    0.154
z_cngew          -0.084744   0.115287  -0.735    0.464
z_son5mon         0.144317   0.111187   1.298    0.198
---
Signif. codes:  0 ‘***’ 0.001 ‘**’ 0.01 ‘*’ 0.05 ‘.’ 0.1 ‘ ’ 1

Residual standard error: 0.9878 on 79 degrees of freedom
Multiple R-squared:  0.09317, Adjusted R-squared:  0.02429
F-statistic: 1.353 on 6 and 79 DF,  p-value: 0.2441

> cat("\n========== MODEL 3b: z_logMeth -> z_son5mot ==========\n")

========== MODEL 3b: z_logMeth -> z_son5mot ==========
> summary(model3_mot)

Call:
lm(formula = z_son5mot ~ z_logMeth + cnsex + Europe_vs_Others +
    z_h3isei + z_cngew + z_son5mon, data = data_complete)

Residuals:
    Min      1Q  Median      3Q     Max
-1.0302 -0.6210 -0.4048  0.3821  3.3913

Coefficients:
                 Estimate Std. Error t value Pr(>|t|)
(Intercept)       0.17600    0.24114   0.730   0.4676
z_logMeth         0.05195    0.10911   0.476   0.6353
cnsex            -0.06972    0.22323  -0.312   0.7556
Europe_vs_Others -0.19417    0.25100  -0.774   0.4415
z_h3isei         -0.21169    0.11190  -1.892   0.0622 .
z_cngew           0.09326    0.11648   0.801   0.4257
z_son5mon        -0.02769    0.11234  -0.246   0.8060
---
```

```
Signif. codes:  0 ‘***’ 0.001 ‘**’ 0.01 ‘*’ 0.05 ‘.’ 0.1 ‘ ’ 1

Residual standard error: 0.998 on 79 degrees of freedom
Multiple R-squared:  0.07434,	Adjusted R-squared:  0.004042 
F-statistic: 1.057 on 6 and 79 DF,  p-value: 0.3952

> cat("\n========== MODEL 3c: z_logMeth -> z_son5koo ==========\n")

========== MODEL 3c: z_logMeth -> z_son5koo ==========
> summary(model3_koo)

Call:
lm(formula = z_son5koo ~ z_logMeth + cnsex + Europe_vs_Others + 
    z_h3isei + z_cngew + z_son5mon, data = data_complete)

Residuals:
    Min      1Q  Median      3Q     Max 
-1.0329 -0.5431 -0.3307  0.0773  3.8672 

Coefficients:
                 Estimate Std. Error t value Pr(>|t|)  
(Intercept)       0.21181    0.23871   0.887   0.3776  
z_logMeth         0.01046    0.10801   0.097   0.9231  
cnsex            -0.25928    0.22098  -1.173   0.2442  
Europe_vs_Others -0.08752    0.24847  -0.352   0.7256  
z_h3isei         -0.22652    0.11078  -2.045   0.0442 *
z_cngew           0.05802    0.11530   0.503   0.6162  
z_son5mon         0.15556    0.11120   1.399   0.1658  
---
Signif. codes:  0 ‘***’ 0.001 ‘**’ 0.01 ‘*’ 0.05 ‘.’ 0.1 ‘ ’ 1

Residual standard error: 0.9879 on 79 degrees of freedom
Multiple R-squared:  0.09289,	Adjusted R-squared:  0.024 
F-statistic: 1.348 on 6 and 79 DF,  p-value: 0.246

> 
> # ============================================================
> # MODEL 4: cngrup -> z_logMeth
> # Covariates: cnsex, Europe_vs_Others, z_h3isei, z_cngew, z_h3tage,
z_cnrandalt, cnerr
> # ============================================================
> 
> model4 <- lm(z_logMeth ~ cngrup + cnsex + Europe_vs_Others + z_h3isei +
z_cngew + 
+                z_h3tage + z_cnrandalt + cnerr,
+              data = data_complete)
> 
> cat("\n========== MODEL 4 ==========\n")

========== MODEL 4 ==========
> summary(model4)

Call:
lm(formula = z_logMeth ~ cngrup + cnsex + Europe_vs_Others + 
    z_h3isei + z_cngew + z_h3tage + z_cnrandalt + cnerr, data =
data_complete)

Residuals:
    Min      1Q  Median      3Q     Max 
-1.2036 -0.6074 -0.3106  0.2909  3.3237 

Coefficients:
                 Estimate Std. Error t value Pr(>|t|)
(Intercept)      -0.49476    0.35699  -1.386   0.1698
```

```
cngrup             0.01464    0.22864   0.064   0.9491
cnsex              0.06315    0.22113   0.286   0.7759
Europe_vs_Others  0.17464    0.24302   0.719   0.4745
z_h3isei         -0.06957    0.11089  -0.627   0.5323
z_cngew            0.05322    0.11460   0.464   0.6437
z_h3tage         -0.20989    0.10982  -1.911   0.0597 .
z_cnrandalt        0.29244    0.11194   2.612   0.0108 *
cnerr              0.38664    0.31441   1.230   0.2225
---
Signif. codes:  0 '***' 0.001 '**' 0.01 '*' 0.05 '.' 0.1 ' ' 1

Residual standard error: 0.9764 on 77 degrees of freedom
Multiple R-squared:  0.1363,   Adjusted R-squared:  0.04655
F-statistic: 1.519 on 8 and 77 DF,  p-value: 0.1646

>
> # ================================================================
> # MODEL 5: Mediation cngrup -> z_logMeth -> z_son5kon
> # ================================================================
>
> med_model5 <- lm(z_logMeth ~ cngrup + cnerr + z_cnrandalt + z_h3tage +
+                     cnsex + Europe_vs_Others + z_cngew + z_h3isei,
+                   data = data_complete)
>
> out_model5 <- lm(z_son5kon ~ cngrup + z_logMeth + cnerr + z_cnrandalt +
+                     cnsex + z_son5mon + Europe_vs_Others + z_h3isei +
z_cngew,
+                   data = data_complete)
>
> set.seed(2026)
> med5 <- mediate(med_model5, out_model5,
+                  treat = "cngrup",
+                  mediator = "z_logMeth",
+                  sims = 1000,
+                  boot = TRUE)
Running nonparametric bootstrap>
> cat("\n========== MODEL 5: cngrup -> z_logMeth -> z_son5kon ==========\n")

========== MODEL 5: cngrup -> z_logMeth -> z_son5kon ==========
> summary(med5)

Causal Mediation Analysis

Nonparametric Bootstrap Confidence Intervals with the Percentile Method

                 Estimate 95% CI Lower 95% CI Upper p-value
ACME            0.0038554   -0.1422266    0.1615643   0.876
ADE             0.0382444   -0.3554379    0.4118113   0.806
Total Effect    0.0420998   -0.3516585    0.4534384   0.808
Prop. Mediated  0.0915783   -4.6580921    6.1462935   0.708

Sample Size Used: 86


Simulations: 1000

>
> # ================================================================
> # MODEL 6: Mediation z_logMeth -> z_son5kon -> z_son5iq
> # ================================================================
>
> med_model6 <- lm(z_son5kon ~ z_logMeth + z_h3tage + cnsex +
+                     Europe_vs_Others + z_cngew + z_h3isei + z_son5mon,
+                   data = data_complete)
```

```
>
> out_model6 <- lm(z_son5iq ~ z_logMeth + z_son5kon + cnsex +
+                      Europe_vs_Others + z_h3isei + z_cngew,
+                    data = data_complete)
>
> set.seed(2026)
> med6 <- mediate(med_model6, out_model6,
+                 treat = "z_logMeth",
+                 mediator = "z_son5kon",
+                 sims = 1000,
+                 boot = TRUE)
Running nonparametric bootstrap>
> cat("\n========== MODEL 6: z_logMeth -> z_son5kon -> z_son5iq
==========\n")

========== MODEL 6: z_logMeth -> z_son5kon -> z_son5iq ==========
> summary(med6)

Causal Mediation Analysis

Nonparametric Bootstrap Confidence Intervals with the Percentile Method

                  Estimate 95% CI Lower 95% CI Upper p-value
ACME            -0.0986658   -0.2215459   -0.0187365   0.012 *
ADE              0.0037977   -0.1647921    0.2478016   0.890
Total Effect    -0.0948680   -0.2646672    0.1376257   0.396
Prop. Mediated   1.0400317  -13.7935244    9.3821351   0.400
---
Signif. codes:  0 ‘***’ 0.001 ‘**’ 0.01 ‘*’ 0.05 ‘.’ 0.1 ‘ ’ 1

Sample Size Used: 86


Simulations: 1000

>
> # ================================================================
> # MODEL 7: Mediation z_e3part_rev -> z_logMeth -> z_son5kon
> # ================================================================
>
> med_model7 <- lm(z_logMeth ~ z_e3part_rev + z_h3tage + cnsex +
+                      Europe_vs_Others + z_cngew + z_h3isei,
+                    data = data_complete)
>
> out_model7 <- lm(z_son5kon ~ z_e3part_rev + z_logMeth + cnsex +
+                      z_son5mon + Europe_vs_Others + z_h3isei + z_cngew,
+                    data = data_complete)
>
> set.seed(2026)
> med7 <- mediate(med_model7, out_model7,
+                 treat = "z_e3part_rev",
+                 mediator = "z_logMeth",
+                 sims = 1000,
+                 boot = TRUE)
Running nonparametric bootstrap>
> cat("\n========== MODEL 7: z_e3part_rev -> z_logMeth -> z_son5kon
==========\n")

========== MODEL 7: z_e3part_rev -> z_logMeth -> z_son5kon ==========
> summary(med7)

Causal Mediation Analysis

Nonparametric Bootstrap Confidence Intervals with the Percentile Method
```

```
                 Estimate 95% CI Lower 95% CI Upper p-value
ACME            0.0759067   -0.0065032    0.2154302   0.076 .
ADE            -0.0142656   -0.2251060    0.1583064   0.878
Total Effect    0.0616411   -0.1426862    0.2515811   0.492
Prop. Mediated  1.2314307   -7.0321355   10.1674359   0.468
---
Signif. codes:  0 ‘***’ 0.001 ‘**’ 0.01 ‘*’ 0.05 ‘.’ 0.1 ‘ ’ 1

Sample Size Used: 86


Simulations: 1000

>
> # ================================================================
> # MODEL 8: Moderated Mediation (separate groups)
> # ================================================================
>
> # Control Group (cngrup = 0)
> cg_data <- data_complete[data_complete$cngrup == 0, ]
> ig_data <- data_complete[data_complete$cngrup == 1, ]
>
> cat("\n========== MODEL 8: Moderated Mediation ==========\n")

========== MODEL 8: Moderated Mediation ==========
> cat("Control Group N:", nrow(cg_data), "\n")
Control Group N: 39
> cat("Intervention Group N:", nrow(ig_data), "\n")
Intervention Group N: 47
>
> # Control Group mediation
> if(nrow(cg_data) >= 20) {
+   med_cg <- lm(z_logMeth ~ z_e3part_rev + z_h3tage + cnsex +
+                  Europe_vs_Others + z_cngew + z_h3isei,
+                data = cg_data)
+   out_cg <- lm(z_son5kon ~ z_e3part_rev + z_logMeth + cnsex +
+                  z_son5mon + Europe_vs_Others + z_h3isei + z_cngew,
+                data = cg_data)
+
+   set.seed(2026)
+   med8_cg <- mediate(med_cg, out_cg,
+                      treat = "z_e3part_rev",
+                      mediator = "z_logMeth",
+                      sims = 1000,
+                      boot = TRUE)
+   cat("\nControl Group:\n")
+   print(summary(med8_cg))
+ }
Running nonparametric bootstrap
Control Group:

Causal Mediation Analysis

Nonparametric Bootstrap Confidence Intervals with the Percentile Method

                Estimate 95% CI Lower 95% CI Upper p-value
ACME            0.127805    -0.054882     0.422445   0.358
ADE            -0.068779    -0.604624     0.388801   0.692
Total Effect    0.059026    -0.529532     0.503156   0.856
Prop. Mediated  2.165240    -5.532001     6.843259   0.678

Sample Size Used: 39
```

```
Simulations: 1000

>
> # Intervention Group mediation
> if(nrow(ig_data) >= 20) {
+   med_ig <- lm(z_logMeth ~ z_e3part_rev + z_h3tage + cnsex +
+                  Europe_vs_Others + z_cngew + z_h3isei,
+                data = ig_data)
+   out_ig <- lm(z_son5kon ~ z_e3part_rev + z_logMeth + cnsex +
+                  z_son5mon + Europe_vs_Others + z_h3isei + z_cngew,
+                data = ig_data)
+
+   set.seed(2026)
+   med8_ig <- mediate(med_ig, out_ig,
+                      treat = "z_e3part_rev",
+                      mediator = "z_logMeth",
+                      sims = 1000,
+                      boot = TRUE)
+   cat("\nIntervention Group:\n")
+   print(summary(med8_ig))
+ }
Running nonparametric bootstrap
Intervention Group:

Causal Mediation Analysis

Nonparametric Bootstrap Confidence Intervals with the Percentile Method

                Estimate 95% CI Lower 95% CI Upper p-value
ACME            0.054892    -0.065009     0.196724   0.364
ADE            -0.021212    -0.279439     0.210536   0.998
Total Effect    0.033681    -0.186221     0.259134   0.720
Prop. Mediated  1.629791    -6.973615     7.920586   0.788

Sample Size Used: 47


Simulations: 1000

>
> # ============================================================
> # MODEL 9: Serial Mediation
> # z_e3part_rev -> z_logMeth -> z_son5kon -> z_son5iq
> # ============================================================
>
> cat("\n========== MODEL 9: Serial Mediation ==========\n")

========== MODEL 9: Serial Mediation ==========
>
> # Path a: z_e3part_rev -> z_logMeth
> model_a <- lm(z_logMeth ~ z_e3part_rev + cnsex + z_h3tage +
+                 Europe_vs_Others + z_cngew + z_h3isei,
+               data = data_complete)
> a_coef <- coef(model_a)["z_e3part_rev"]
>
> # Path b1: z_logMeth -> z_son5kon
> model_b1 <- lm(z_son5kon ~ z_logMeth + cnsex + z_son5mon +
+                  Europe_vs_Others + z_cngew + z_h3isei,
+                data = data_complete)
> b1_coef <- coef(model_b1)["z_logMeth"]
>
> # Path b2: z_son5kon -> z_son5iq
```

```
> model_b2 <- lm(z_son5iq ~ z_son5kon + cnsex + Europe_vs_Others + z_cngew +
z_h3isei,
+                data = data_complete)
> b2_coef <- coef(model_b2)["z_son5kon"]
>
> # Indirect effects
> ab1 <- a_coef * b1_coef
> ab2 <- b1_coef * b2_coef
> ab_serial <- a_coef * b1_coef * b2_coef
> total_indirect <- ab1 + ab2 + ab_serial
>
> # Total effect
> model_total <- lm(z_son5iq ~ z_e3part_rev + cnsex + Europe_vs_Others +
z_cngew + z_h3isei,
+                   data = data_complete)
> total_effect <- coef(model_total)["z_e3part_rev"]
>
> # Direct effect (c')
> model_cprime <- lm(z_son5iq ~ z_e3part_rev + z_logMeth + z_son5kon +
+                      cnsex + Europe_vs_Others + z_cngew + z_h3isei,
+                    data = data_complete)
> cprime_coef <- coef(model_cprime)["z_e3part_rev"]
>
> # Bootstrap for serial indirect effect
> boot_serial <- function(data, indices) {
+   d <- data[indices, ]
+   a_boot <- coef(lm(z_logMeth ~ z_e3part_rev + cnsex + z_h3tage +
+                       Europe_vs_Others + z_cngew + z_h3isei, data =
d))["z_e3part_rev"]
+   b1_boot <- coef(lm(z_son5kon ~ z_logMeth + cnsex + z_son5mon +
+                        Europe_vs_Others + z_cngew + z_h3isei, data =
d))["z_logMeth"]
+   b2_boot <- coef(lm(z_son5iq ~ z_son5kon + cnsex + Europe_vs_Others +
+                        z_cngew + z_h3isei, data = d))["z_son5kon"]
+   return(a_boot * b1_boot * b2_boot)
+ }
>
> set.seed(2026)
> boot_result <- boot::boot(data_complete, boot_serial, R = 1000)
> boot_se <- sd(boot_result$t, na.rm = TRUE)
> boot_ci <- quantile(boot_result$t, c(0.025, 0.975), na.rm = TRUE)
> p_serial <- 2 * (1 - pnorm(abs(ab_serial / boot_se)))
>
> cat("\nSerial Mediation Results:\n")

Serial Mediation Results:
> cat("  a (z_e3part_rev -> z_logMeth):", round(a_coef, 4), "\n")
  a (z_e3part_rev -> z_logMeth): 0.2388
> cat("  b1 (z_logMeth -> z_son5kon):", round(b1_coef, 4), "\n")
  b1 (z_logMeth -> z_son5kon): 0.3145
> cat("  b2 (z_son5kon -> z_son5iq):", round(b2_coef, 4), "\n")
  b2 (z_son5kon -> z_son5iq): -0.3605
> cat("  Indirect via methylation only (ab1):", round(ab1, 4), "\n")
  Indirect via methylation only (ab1): 0.0751
> cat("  Indirect via concentration only (ab2):", round(ab2, 4), "\n")
  Indirect via concentration only (ab2): -0.1134
> cat("  Serial indirect (a*b1*b2):", round(ab_serial, 4), "\n")
  Serial indirect (a*b1*b2): -0.0271
> cat("  Boot SE:", round(boot_se, 4), "\n")
  Boot SE: 0.0234
> cat("  Boot 95% CI: [", round(boot_ci[1], 4), ",", round(boot_ci[2], 4),
"]\n")
  Boot 95% CI: [ -0.0856 , 0.0023 ]
> cat("  p-value:", round(p_serial, 4), "\n")
```

```
  p-value: 0.2477
> cat("  Total indirect:", round(total_indirect, 4), "\n")
  Total indirect: -0.0654
> cat("  Direct effect (c'):", round(cprime_coef, 4), "\n")
  Direct effect (c'): -0.0287
> cat("  Total effect:", round(total_effect, 4), "\n")
  Total effect: -0.045
>
> # ============================================================
> # MODEL 10: Moderated Serial Mediation (IG vs CG)
> # ============================================================
>
> cat("\n========== MODEL 10: Moderated Serial Mediation ==========\n")

========== MODEL 10: Moderated Serial Mediation ==========
>
> run_serial_group <- function(data, group_name) {
+
+   cat("\n", group_name, "N =", nrow(data), "\n")
+
+   if(nrow(data) < 20) {
+     return(list(ab_serial = NA, p = NA, n = nrow(data)))
+   }
+
+   # Paths
+   model_a <- lm(z_logMeth ~ z_e3part_rev + cnsex + z_h3tage +
+                   Europe_vs_Others + z_cngew + z_h3isei, data = data)
+   a_coef <- coef(model_a)["z_e3part_rev"]
+
+   model_b1 <- lm(z_son5kon ~ z_logMeth + cnsex + z_son5mon +
+                    Europe_vs_Others + z_cngew + z_h3isei, data = data)
+   b1_coef <- coef(model_b1)["z_logMeth"]
+
+   model_b2 <- lm(z_son5iq ~ z_son5kon + cnsex + Europe_vs_Others + z_cngew
+ z_h3isei,
+                  data = data)
+   b2_coef <- coef(model_b2)["z_son5kon"]
+
+   ab_serial <- a_coef * b1_coef * b2_coef
+
+   # Bootstrap
+   boot_serial <- function(d, indices) {
+     d_sub <- d[indices, ]
+     a_boot <- coef(lm(z_logMeth ~ z_e3part_rev + cnsex + z_h3tage +
+                         Europe_vs_Others + z_cngew + z_h3isei, data =
d_sub))["z_e3part_rev"]
+     b1_boot <- coef(lm(z_son5kon ~ z_logMeth + cnsex + z_son5mon +
+                          Europe_vs_Others + z_cngew + z_h3isei, data =
d_sub))["z_logMeth"]
+     b2_boot <- coef(lm(z_son5iq ~ z_son5kon + cnsex + Europe_vs_Others +
+                          z_cngew + z_h3isei, data = d_sub))["z_son5kon"]
+     return(a_boot * b1_boot * b2_boot)
+   }
+
+   set.seed(2026)
+   boot_result <- boot::boot(data, boot_serial, R = 500)
+   boot_se <- sd(boot_result$t, na.rm = TRUE)
+   p_serial <- 2 * (1 - pnorm(abs(ab_serial / boot_se)))
+
+   return(list(ab_serial = ab_serial, p = p_serial, n = nrow(data),
+               a = a_coef, b1 = b1_coef, b2 = b2_coef))
+ }
>
> # Run for both groups
```

```
> cg_serial <- run_serial_group(data_complete[data_complete$cngrup == 0, ],
"Control Group")

 Control Group N = 39
> ig_serial <- run_serial_group(data_complete[data_complete$cngrup == 1, ],
"Intervention Group")

 Intervention Group N = 47
>
> cat("\n--- Results ---\n")

--- Results ---
> cat("Control Group:\n")
Control Group:
> cat("  N:", cg_serial$n, "\n")
  N: 39
> cat("  Serial indirect:", round(cg_serial$ab_serial, 4), "\n")
  Serial indirect: -0.0555
> cat("  p-value:", round(cg_serial$p, 4), "\n\n")
  p-value: 0.384

>
> cat("Intervention Group:\n")
Intervention Group:
> cat("  N:", ig_serial$n, "\n")
  N: 47
> cat("  Serial indirect:", round(ig_serial$ab_serial, 4), "\n")
  Serial indirect: -0.018
> cat("  p-value:", round(ig_serial$p, 4), "\n\n")
  p-value: 0.3736

>
> # Test difference
> if(!is.na(cg_serial$ab_serial) & !is.na(ig_serial$ab_serial)) {
+   diff_serial <- ig_serial$ab_serial - cg_serial$ab_serial
+   cat("Difference (IG - CG):", round(diff_serial, 4), "\n")
+ }
Difference (IG - CG): 0.0375
```

**NMAR analyses**

```
> library(dplyr)
> library(mediation)
> library(boot)
> # Load raw data
> raw_data <- read.csv("original dataset.csv")
> dummy_codes <- c(-333, -444, -555, -777, -999)
> data_clean <- raw_data
> for(code in dummy_codes) {
+   data_clean[data_clean == code] <- NA
+ }
> data_selected <- data_clean %>%
+   dplyr::select(cngrup, cnsex, h3tage, cnrandalt, h3isei, cngew,
+                 Europe_vs_Others, cnerr, logMeth, e3part,
+                 son5iq, son5kon, son5ver, son5mot, son5koo, son5mon)
> missing_h3isei <- is.na(data_selected$h3isei)
> cat("Missing on h3isei:", sum(missing_h3isei),
+     "(", round(mean(missing_h3isei)*100, 1), "%)\n")
Missing on h3isei: 16 ( 12.1 %)
> set.seed(2026)
> observed_h3isei <- data_selected$h3isei[!missing_h3isei]
```

```
> mean_h3isei <- mean(observed_h3isei, na.rm = TRUE)
> sd_h3isei <- sd(observed_h3isei, na.rm = TRUE)
> cat("\nObserved h3isei - Mean:", round(mean_h3isei, 3),
+     "SD:", round(sd_h3isei, 3), "\n")

Observed h3isei - Mean: 34.44 SD: 23.206
> # Shift -1 SD (lower values for missing)
> shift_h3isei <- -1
> imputed_h3isei <- mean_h3isei + shift_h3isei * sd_h3isei +
+   rnorm(sum(missing_h3isei), 0, sd_h3isei * 0.3)
> cat("Imputed h3isei (shift -1 SD) - Mean:", round(mean(imputed_h3isei), 3),
+     "SD:", round(sd(imputed_h3isei), 3), "\n")
Imputed h3isei (shift -1 SD) - Mean: 7.497 SD: 6.856
> data_mnar <- data_selected
> # Impute h3isei (shifted down)
> data_mnar$h3isei[missing_h3isei] <- imputed_h3isei
> # For other variables, use median imputation (MAR)
> impute_with_median <- function(x) {
+   ifelse(is.na(x), median(x, na.rm = TRUE), x)
+ }
> numeric_vars <- sapply(data_mnar, is.numeric)
> data_mnar[, numeric_vars] <- lapply(data_mnar[, numeric_vars],
impute_with_median)
> # Remove any remaining NA
> data_complete <- na.omit(data_mnar)
> cat("\nFinal MNAR dataset N (Models 1-6):", nrow(data_complete), "\n")

Final MNAR dataset N (Models 1-6): 132
> cat("Shift applied: h3isei -1 SD\n")
Shift applied: h3isei -1 SD
> data_complete <- data_complete %>%
+   mutate(
+     z_logMeth = as.numeric(scale(logMeth)),
+     z_h3isei = as.numeric(scale(h3isei)),
+     z_cngew = as.numeric(scale(cngew)),
+     z_h3tage = as.numeric(scale(h3tage)),
+     z_cnrandalt = as.numeric(scale(cnrandalt)),
+     z_son5iq = as.numeric(scale(son5iq)),
+     z_son5kon = as.numeric(scale(son5kon)),
+     z_son5ver = as.numeric(scale(son5ver)),
+     z_son5mot = as.numeric(scale(son5mot)),
+     z_son5koo = as.numeric(scale(son5koo)),
+     z_son5mon = as.numeric(scale(son5mon)),
+     z_e3part = as.numeric(scale(e3part))
+   )
> cat("\nZ-scores created.\n")

Z-scores created.
> model1 <- lm(z_son5iq ~ z_logMeth + cnsex + Europe_vs_Others + z_h3isei +
z_cngew,
+              data = data_complete)
> cat("\n========== MODEL 1 (MNAR, h3isei -1 SD) ==========\n")

========== MODEL 1 (MNAR, h3isei -1 SD) ==========
> summary(model1)

Call:
lm(formula = z_son5iq ~ z_logMeth + cnsex + Europe_vs_Others +
    z_h3isei + z_cngew, data = data_complete)

Residuals:
     Min       1Q   Median       3Q      Max
-2.47479 -0.50615 -0.00185  0.59697  2.18711
```

```
Coefficients:
                 Estimate Std. Error t value Pr(>|t|)
(Intercept)      -0.08975    0.18431  -0.487  0.62715
z_logMeth        -0.07957    0.08406  -0.947  0.34569
cnsex             0.30212    0.16897   1.788  0.07619 .
Europe_vs_Others -0.10538    0.19225  -0.548  0.58454
z_h3isei          0.23026    0.08625   2.670  0.00859 **
z_cngew           0.19330    0.08583   2.252  0.02604 *
---
Signif. codes:  0 ‘***’ 0.001 ‘**’ 0.01 ‘*’ 0.05 ‘.’ 0.1 ‘ ’ 1

Residual standard error: 0.9594 on 126 degrees of freedom
Multiple R-squared:  0.1147,	Adjusted R-squared:  0.07953
F-statistic: 3.264 on 5 and 126 DF,  p-value: 0.008339

> model2 <- lm(z_son5kon ~ z_logMeth + cnsex + Europe_vs_Others + z_h3isei +
z_cngew + z_son5mon,
+              data = data_complete)
> cat("\n========== MODEL 2 (MNAR, h3isei -1 SD) ==========\n")

========== MODEL 2 (MNAR, h3isei -1 SD) ==========
> summary(model2)

Call:
lm(formula = z_son5kon ~ z_logMeth + cnsex + Europe_vs_Others +
    z_h3isei + z_cngew + z_son5mon, data = data_complete)

Residuals:
    Min      1Q  Median      3Q     Max
-1.5858 -0.8407  0.1452  0.5663  3.1910

Coefficients:
                  Estimate Std. Error t value Pr(>|t|)
(Intercept)       0.191795   0.190135   1.009  0.31505
z_logMeth         0.225264   0.085357   2.639  0.00937 **
cnsex            -0.377073   0.173897  -2.168  0.03202 *
Europe_vs_Others  0.019493   0.196807   0.099  0.92126
z_h3isei         -0.122945   0.087795  -1.400  0.16388
z_cngew           0.028394   0.088760   0.320  0.74958
z_son5mon        -0.003638   0.088195  -0.041  0.96716
---
Signif. codes:  0 ‘***’ 0.001 ‘**’ 0.01 ‘*’ 0.05 ‘.’ 0.1 ‘ ’ 1

Residual standard error: 0.9737 on 125 degrees of freedom
Multiple R-squared:  0.09531,	Adjusted R-squared:  0.05189
F-statistic: 2.195 on 6 and 125 DF,  p-value: 0.04774

> model3_ver <- lm(z_son5ver ~ z_logMeth + cnsex + Europe_vs_Others +
z_h3isei + z_cngew + z_son5mon,
+                  data = data_complete)
> model3_mot <- lm(z_son5mot ~ z_logMeth + cnsex + Europe_vs_Others +
z_h3isei + z_cngew + z_son5mon,
+                  data = data_complete)
> model3_koo <- lm(z_son5koo ~ z_logMeth + cnsex + Europe_vs_Others +
z_h3isei + z_cngew + z_son5mon,
+                  data = data_complete)
> cat("\n========== MODEL 3a (MNAR): z_logMeth -> z_son5ver ==========\n")

========== MODEL 3a (MNAR): z_logMeth -> z_son5ver ==========
> summary(model3_ver)

Call:
lm(formula = z_son5ver ~ z_logMeth + cnsex + Europe_vs_Others +
    z_h3isei + z_cngew + z_son5mon, data = data_complete)
```

```
Residuals:
    Min      1Q  Median      3Q     Max 
-1.1951 -0.3914 -0.2643 -0.0912  3.9511 

Coefficients:
                 Estimate Std. Error t value Pr(>|t|)  
(Intercept)       0.09293    0.19371   0.480   0.6323  
z_logMeth         0.18296    0.08696   2.104   0.0374 *
cnsex            -0.07568    0.17717  -0.427   0.6700  
Europe_vs_Others -0.07253    0.20051  -0.362   0.7182  
z_h3isei         -0.11968    0.08945  -1.338   0.1833  
z_cngew          -0.07303    0.09043  -0.808   0.4209  
z_son5mon         0.07478    0.08985   0.832   0.4068  
---
Signif. codes:  0 ‘***’ 0.001 ‘**’ 0.01 ‘*’ 0.05 ‘.’ 0.1 ‘ ’ 1

Residual standard error: 0.992 on 125 degrees of freedom
Multiple R-squared:  0.06097,	Adjusted R-squared:  0.01589 
F-statistic: 1.353 on 6 and 125 DF,  p-value: 0.239

> cat("\n========== MODEL 3b (MNAR): z_logMeth -> z_son5mot ==========\n")

========== MODEL 3b (MNAR): z_logMeth -> z_son5mot ==========
> summary(model3_mot)

Call:
lm(formula = z_son5mot ~ z_logMeth + cnsex + Europe_vs_Others + 
    z_h3isei + z_cngew + z_son5mon, data = data_complete)

Residuals:
    Min      1Q  Median      3Q     Max 
-0.9581 -0.6741 -0.4114  0.5872  3.5299 

Coefficients:
                 Estimate Std. Error t value Pr(>|t|)  
(Intercept)       0.20762    0.19319   1.075   0.2846  
z_logMeth         0.06863    0.08673   0.791   0.4303  
cnsex            -0.11072    0.17670  -0.627   0.5321  
Europe_vs_Others -0.20675    0.19997  -1.034   0.3032  
z_h3isei         -0.20561    0.08921  -2.305   0.0228 *
z_cngew          -0.03758    0.09019  -0.417   0.6776  
z_son5mon        -0.03429    0.08961  -0.383   0.7026  
---
Signif. codes:  0 ‘***’ 0.001 ‘**’ 0.01 ‘*’ 0.05 ‘.’ 0.1 ‘ ’ 1

Residual standard error: 0.9894 on 125 degrees of freedom
Multiple R-squared:  0.06596,	Adjusted R-squared:  0.02113 
F-statistic: 1.471 on 6 and 125 DF,  p-value: 0.1933

> cat("\n========== MODEL 3c (MNAR): z_logMeth -> z_son5koo ==========\n")

========== MODEL 3c (MNAR): z_logMeth -> z_son5koo ==========
> summary(model3_koo)

Call:
lm(formula = z_son5koo ~ z_logMeth + cnsex + Europe_vs_Others + 
    z_h3isei + z_cngew + z_son5mon, data = data_complete)

Residuals:
    Min      1Q  Median      3Q     Max 
-0.9346 -0.4605 -0.2973 -0.0904  4.7858 

Coefficients:
```

```
                   Estimate Std. Error t value Pr(>|t|)
(Intercept)       0.2784702  0.1944822   1.432    0.155
z_logMeth         0.0093324  0.0873082   0.107    0.915
cnsex            -0.1366084  0.1778733  -0.768    0.444
Europe_vs_Others -0.2864070  0.2013073  -1.423    0.157
z_h3isei         -0.1394891  0.0898022  -1.553    0.123
z_cngew           0.0002158  0.0907898   0.002    0.998
z_son5mon         0.0558172  0.0902111   0.619    0.537

Residual standard error: 0.996 on 125 degrees of freedom
Multiple R-squared:  0.05347,	Adjusted R-squared:  0.008034 
F-statistic: 1.177 on 6 and 125 DF,  p-value: 0.3229

> model4 <- lm(z_logMeth ~ cngrup + cnsex + Europe_vs_Others + z_h3isei + 
z_cngew +
+                z_h3tage + z_cnrandalt + cnerr,
+              data = data_complete)
> cat("\n========== MODEL 4 (MNAR, h3isei -1 SD) ==========\n")

========== MODEL 4 (MNAR, h3isei -1 SD) ==========
> summary(model4)

Call:
lm(formula = z_logMeth ~ cngrup + cnsex + Europe_vs_Others + 
    z_h3isei + z_cngew + z_h3tage + z_cnrandalt + cnerr, data = 
data_complete)

Residuals:
    Min      1Q  Median      3Q     Max 
-1.2317 -0.6131 -0.2772  0.2390  3.6965 

Coefficients:
                 Estimate Std. Error t value Pr(>|t|)   
(Intercept)      -0.33216    0.30616  -1.085  0.28007   
cngrup           -0.36692    0.17958  -2.043  0.04317 * 
cnsex             0.15506    0.17313   0.896  0.37221   
Europe_vs_Others  0.06878    0.20022   0.344  0.73178   
z_h3isei         -0.04598    0.08952  -0.514  0.60839   
z_cngew          -0.04346    0.08861  -0.490  0.62467   
z_h3tage         -0.13840    0.08909  -1.553  0.12288   
z_cnrandalt       0.23504    0.08951   2.626  0.00974 **
cnerr             0.45374    0.26831   1.691  0.09335 . 
---
Signif. codes:  0 ‘***’ 0.001 ‘**’ 0.01 ‘*’ 0.05 ‘.’ 0.1 ‘ ’ 1

Residual standard error: 0.9745 on 123 degrees of freedom
Multiple R-squared:  0.1084,	Adjusted R-squared:  0.05043 
F-statistic:  1.87 on 8 and 123 DF,  p-value: 0.07067

> med_model5 <- lm(z_logMeth ~ cngrup + cnerr + z_cnrandalt + z_h3tage +
+                    cnsex + Europe_vs_Others + z_cngew + z_h3isei,
+                  data = data_complete)
> out_model5 <- lm(z_son5kon ~ cngrup + z_logMeth + cnerr + z_cnrandalt +
+                    cnsex + z_son5mon + Europe_vs_Others + z_h3isei + 
z_cngew,
+                  data = data_complete)
> set.seed(2026)
> med5 <- mediate(med_model5, out_model5,
+                 treat = "cngrup",
+                 mediator = "z_logMeth",
+                 sims = 1000,
+                 boot = TRUE)
Running nonparametric bootstrap> cat("\n========== MODEL 5 (MNAR): cngrup -> 
z_logMeth -> z_son5kon ==========\n")
```

```
========== MODEL 5 (MNAR): cngrup -> z_logMeth -> z_son5kon ==========
> summary(med5)

Causal Mediation Analysis

Nonparametric Bootstrap Confidence Intervals with the Percentile Method

                Estimate 95% CI Lower 95% CI Upper p-value
ACME           -0.076318    -0.190301     0.011892   0.110
ADE             0.028458    -0.373995     0.376441   0.864
Total Effect   -0.047861    -0.439014     0.314359   0.806
Prop. Mediated  1.594594    -7.533051     6.723051   0.820

Sample Size Used: 132


Simulations: 1000

> med_model6 <- lm(z_son5kon ~ z_logMeth + z_h3tage + cnsex +
+                    Europe_vs_Others + z_cngew + z_h3isei + z_son5mon,
+                  data = data_complete)
> out_model6 <- lm(z_son5iq ~ z_logMeth + z_son5kon + cnsex +
+                    Europe_vs_Others + z_h3isei + z_cngew,
+                  data = data_complete)
> set.seed(2026)
> med6 <- mediate(med_model6, out_model6,
+                 treat = "z_logMeth",
+                 mediator = "z_son5kon",
+                 sims = 1000,
+                 boot = TRUE)
Running nonparametric bootstrap> cat("\n========== MODEL 6 (MNAR): z_logMeth
-> z_son5kon -> z_son5iq ==========\n")

========== MODEL 6 (MNAR): z_logMeth -> z_son5kon -> z_son5iq ==========
> summary(med6)

Causal Mediation Analysis

Nonparametric Bootstrap Confidence Intervals with the Percentile Method

                 Estimate 95% CI Lower 95% CI Upper p-value
ACME           -0.0780908   -0.1561031   -0.0096382   0.032 *
ADE             0.0029809   -0.1451244    0.1926256   0.932
Total Effect   -0.0751098   -0.2275144    0.1129509   0.430
Prop. Mediated  1.0396878  -10.9486968    8.5211390   0.426
---
Signif. codes:  0 ‘***’ 0.001 ‘**’ 0.01 ‘*’ 0.05 ‘.’ 0.1 ‘ ’ 1

Sample Size Used: 132


Simulations: 1000

> med_model7 <- lm(z_logMeth ~ z_e3part_rev + z_h3tage + cnsex +
+                    Europe_vs_Others + z_cngew + z_h3isei,
+                  data = data_complete)
> out_model7 <- lm(z_son5kon ~ z_e3part_rev + z_logMeth + cnsex +
+                    z_son5mon + Europe_vs_Others + z_h3isei + z_cngew,
+                  data = data_complete)
> set.seed(2026)
> med7 <- mediate(med_model7, out_model7,
+                 treat = "z_e3part_rev",
+                 mediator = "z_logMeth",
```

```
+                  sims = 1000,
+                  boot = TRUE)
Running nonparametric bootstrap> cat("\n========== MODEL 7 (MNAR)
==========\n")

========== MODEL 7 (MNAR) ==========
> cat("z_e3part_rev -> z_logMeth -> z_son5kon\n")
z_e3part_rev -> z_logMeth -> z_son5kon
> cat("Shift: e3part_rev +1 SD, h3isei -1 SD\n")
Shift: e3part_rev +1 SD, h3isei -1 SD
> summary(med7)

Causal Mediation Analysis

Nonparametric Bootstrap Confidence Intervals with the Percentile Method

                 Estimate 95% CI Lower 95% CI Upper p-value
ACME            0.0536573    0.0028909    0.1293542   0.032 *
ADE             0.0260346   -0.1675971    0.2008616   0.792
Total Effect    0.0796919   -0.1034261    0.2489432   0.370
Prop. Mediated  0.6733093   -4.1153945    6.8675909   0.374
---
Signif. codes:  0 ‘***’ 0.001 ‘**’ 0.01 ‘*’ 0.05 ‘.’ 0.1 ‘ ’ 1

Sample Size Used: 132


Simulations: 1000

> cg_data <- data_complete[data_complete$cngrup == 0, ]
> ig_data <- data_complete[data_complete$cngrup == 1, ]
> cat("\n========== MODEL 8 (MNAR) ==========\n")

========== MODEL 8 (MNAR) ==========
> cat("Control Group N:", nrow(cg_data), "\n")
Control Group N: 60
> cat("Intervention Group N:", nrow(ig_data), "\n")
Intervention Group N: 72
> if(nrow(cg_data) >= 20) {
+   med_cg <- lm(z_logMeth ~ z_e3part_rev + z_h3tage + cnsex +
+                  Europe_vs_Others + z_cngew + z_h3isei,
+                data = cg_data)
+   out_cg <- lm(z_son5kon ~ z_e3part_rev + z_logMeth + cnsex +
+                  z_son5mon + Europe_vs_Others + z_h3isei + z_cngew,
+                data = cg_data)
+
+   set.seed(2026)
+   med8_cg <- mediate(med_cg, out_cg,
+                      treat = "z_e3part_rev",
+                      mediator = "z_logMeth",
+                      sims = 1000,
+                      boot = TRUE)
+   cat("\nControl Group:\n")
+   print(summary(med8_cg))
+ }
Running nonparametric bootstrap
Control Group:

Causal Mediation Analysis

Nonparametric Bootstrap Confidence Intervals with the Percentile Method

                Estimate 95% CI Lower 95% CI Upper p-value
ACME            0.067069    -0.058259     0.273373   0.292
```

```
ADE               0.081263    -0.298525     0.368472   0.646
Total Effect      0.148331    -0.210913     0.450828   0.336
Prop. Mediated    0.452156    -3.666973     4.525034   0.492

Sample Size Used: 60


Simulations: 1000

> if(nrow(ig_data) >= 20) {
+   med_ig <- lm(z_logMeth ~ z_e3part_rev + z_h3tage + cnsex +
+                  Europe_vs_Others + z_cngew + z_h3isei,
+                data = ig_data)
+   out_ig <- lm(z_son5kon ~ z_e3part_rev + z_logMeth + cnsex +
+                  z_son5mon + Europe_vs_Others + z_h3isei + z_cngew,
+                data = ig_data)
+
+   set.seed(2026)
+   med8_ig <- mediate(med_ig, out_ig,
+                      treat = "z_e3part_rev",
+                      mediator = "z_logMeth",
+                      sims = 1000,
+                      boot = TRUE)
+   cat("\nIntervention Group:\n")
+   print(summary(med8_ig))
+ }
Running nonparametric bootstrap
Intervention Group:

Causal Mediation Analysis

Nonparametric Bootstrap Confidence Intervals with the Percentile Method

                 Estimate 95% CI Lower 95% CI Upper p-value
ACME             0.0289918   -0.0401671    0.1089654   0.360
ADE              0.0038628   -0.2392135    0.2205546   0.882
Total Effect     0.0328546   -0.1839784    0.2458732   0.758
Prop. Mediated   0.8824273   -5.3588417    4.0893249   0.830

Sample Size Used: 72


Simulations: 1000

> cat("\n========== MODEL 9 (MNAR) ==========\n")

========== MODEL 9 (MNAR) ==========
> model_a <- lm(z_logMeth ~ z_e3part_rev + cnsex + z_h3tage +
+                 Europe_vs_Others + z_cngew + z_h3isei,
+               data = data_complete)
> a_coef <- coef(model_a)["z_e3part_rev"]
> model_b1 <- lm(z_son5kon ~ z_logMeth + cnsex + z_son5mon +
+                  Europe_vs_Others + z_cngew + z_h3isei,
+                data = data_complete)
> b1_coef <- coef(model_b1)["z_logMeth"]
> model_b2 <- lm(z_son5iq ~ z_son5kon + cnsex + Europe_vs_Others + z_cngew +
z_h3isei,
+                data = data_complete)
> b2_coef <- coef(model_b2)["z_son5kon"]
> ab_serial <- a_coef * b1_coef * b2_coef
> boot_serial <- function(data, indices) {
+   d <- data[indices, ]
+   a_boot <- coef(lm(z_logMeth ~ z_e3part_rev + cnsex + z_h3tage +
```

```
+                         Europe_vs_Others + z_cngew + z_h3isei, data =
d))["z_e3part_rev"]
+   b1_boot <- coef(lm(z_son5kon ~ z_logMeth + cnsex + z_son5mon +
+                          Europe_vs_Others + z_cngew + z_h3isei, data =
d))["z_logMeth"]
+   b2_boot <- coef(lm(z_son5iq ~ z_son5kon + cnsex + Europe_vs_Others +
+                          z_cngew + z_h3isei, data = d))["z_son5kon"]
+   return(a_boot * b1_boot * b2_boot)
+ }
> set.seed(2026)
> boot_result <- boot::boot(data_complete, boot_serial, R = 1000)
> boot_se <- sd(boot_result$t, na.rm = TRUE)
> boot_ci <- quantile(boot_result$t, c(0.025, 0.975), na.rm = TRUE)
> p_serial <- 2 * (1 - pnorm(abs(ab_serial / boot_se)))
> cat("\nSerial Mediation Results (MNAR):\n")

Serial Mediation Results (MNAR):
> cat("  a:", round(a_coef, 4), "\n")
  a: 0.2463
> cat("  b1:", round(b1_coef, 4), "\n")
  b1: 0.2239
> cat("  b2:", round(b2_coef, 4), "\n")
  b2: -0.3657
> cat("  Serial indirect (a*b1*b2):", round(ab_serial, 4), "\n")
  Serial indirect (a*b1*b2): -0.0202
> cat("  Boot SE:", round(boot_se, 4), "\n")
  Boot SE: 0.0139
> cat("  Boot 95% CI: [", round(boot_ci[1], 4), ",", round(boot_ci[2], 4),
"]\n")
  Boot 95% CI: [ -0.0529 , -0.0013 ]
> cat("  p-value:", round(p_serial, 4), "\n")
  p-value: 0.1462
> cat("\n========== MODEL 10 (MNAR) ==========\n")



========== MODEL 10 (MNAR) ==========
> run_serial_group <- function(data, group_name) {
+
+   cat("\n", group_name, "N =", nrow(data), "\n")
+
+   if(nrow(data) < 20) {
+     return(list(ab_serial = NA, p = NA, n = nrow(data)))
+   }
+
+   model_a <- lm(z_logMeth ~ z_e3part_rev + cnsex + z_h3tage +
+                   Europe_vs_Others + z_cngew + z_h3isei, data = data)
+   a_coef <- coef(model_a)["z_e3part_rev"]
+
+   model_b1 <- lm(z_son5kon ~ z_logMeth + cnsex + z_son5mon +
+                    Europe_vs_Others + z_cngew + z_h3isei, data = data)
+   b1_coef <- coef(model_b1)["z_logMeth"]
+
+   model_b2 <- lm(z_son5iq ~ z_son5kon + cnsex + Europe_vs_Others + z_cngew
+ z_h3isei,
+                  data = data)
+   b2_coef <- coef(model_b2)["z_son5kon"]
+
+   ab_serial <- a_coef * b1_coef * b2_coef
+
+   boot_serial <- function(d, indices) {
+     d_sub <- d[indices, ]
+     a_boot <- coef(lm(z_logMeth ~ z_e3part_rev + cnsex + z_h3tage +
```

```
+                          Europe_vs_Others + z_cngew + z_h3isei, data = 
d_sub))["z_e3part_rev"]
+     b1_boot <- coef(lm(z_son5kon ~ z_logMeth + cnsex + z_son5mon + 
+                           Europe_vs_Others + z_cngew + z_h3isei, data = 
d_sub))["z_logMeth"]
+     b2_boot <- coef(lm(z_son5iq ~ z_son5kon + cnsex + Europe_vs_Others + 
+                           z_cngew + z_h3isei, data = d_sub))["z_son5kon"]
+     return(a_boot * b1_boot * b2_boot)
+   }
+ 
+   set.seed(2026)
+   boot_result <- boot::boot(data, boot_serial, R = 500)
+   boot_se <- sd(boot_result$t, na.rm = TRUE)
+   p_serial <- 2 * (1 - pnorm(abs(ab_serial / boot_se)))
+ 
+   return(list(ab_serial = ab_serial, p = p_serial, n = nrow(data)))
+ }
> cg_serial <- run_serial_group(data_complete[data_complete$cngrup == 0, ], 
"Control Group")

 Control Group N = 39 
> ig_serial <- run_serial_group(data_complete[data_complete$cngrup == 1, ], 
"Intervention Group")

 Intervention Group N = 47 
> cat("\n--- Results (MNAR) ---\n")

--- Results (MNAR) ---
> cat("Control Group:\n")
Control Group:
> cat("  N:", cg_serial$n, "\n")
  N: 39 
> cat("  Serial indirect:", round(cg_serial$ab_serial, 4), "\n")
  Serial indirect: -0.0555 
> cat("  p-value:", round(cg_serial$p, 4), "\n\n")
  p-value: 0.384 

> cat("Intervention Group:\n")
Intervention Group:
> cat("  N:", ig_serial$n, "\n")
  N: 47 
> cat("  Serial indirect:", round(ig_serial$ab_serial, 4), "\n")
  Serial indirect: -0.018 
> cat("  p-value:", round(ig_serial$p, 4), "\n\n")
  p-value: 0.3736 

> if(!is.na(cg_serial$ab_serial) & !is.na(ig_serial$ab_serial)) {
+   diff_serial <- ig_serial$ab_serial - cg_serial$ab_serial
+   cat("Difference (IG - CG):", round(diff_serial, 4), "\n")
+ }
Difference (IG - CG): 0.0375
```
